# Peripheral Metabolic-Redox Signaling as a Core Mechanism of Major Depressive Disorder: Evidence From Deep Metabolomic Phenotyping

**DOI:** 10.64898/2025.12.15.25342323

**Authors:** Michael Maes, Mengqi Niu, Annabel Maes, Yiping Luo, Chenkai Yangyang, Jing Li, Abbas F. Almulla, Yingqian Zhang

## Abstract

**Background:** Major depressive disorder (MDD) is a neuro-immune, oxidative, and nitrosative stress (NIMETOX) disorder, in which peripheral immune-redox pathways intersect with metabolic networks leading to neurotoxicity within the limbic-prefrontal affective circuits. Comprehensive metabolomics analysis in well-phenotyped patients is vital to elucidate their metabolic profile.

**Objectives:** To identify metabolic abnormalities that differentiate inpatients with severe MDD from healthy controls through high-resolution, untargeted metabolomics.

**Methods:** Serum samples from 125 MDD inpatients and 40 healthy controls were analyzed utilizing liquid chromatography and mass spectrometry. A meticulously regulated multistage machine learning pipeline with leakage-prevention protocols was employed to analyze differences between MDD and controls and to predict phenome scores.

**Results:** Feature selection showed that 16 metabolites and 6 functional modules reliably distinguished MDD. The functional profile of the metabolites indicates a convergence of lipotoxicity, phospholipid remodeling, disruptions in fatty acid metabolism, mitochondrial redox imbalance, ether-lipid metabolism, and antioxidant depletion. This MDD metabotype was not affected by metabolic syndrome. A substantial portion of the variance in overall depression severity (72.5%), physiosomatic symptoms (55.8%) and suicidal ideation (23.6%) was accounted for by increased lipitoxicity, phospholipid remodeling, and fatty acid storage/signaling. The recurrence of illness (27.7%) was associated with a self-reinforcing-lipid-redox-inflammatory module that maintains cellular stress.

**Discussion:** The MDD metabotype represents a cohesive metabolic network that is associated with the NIMETOX pathogenesis of MDD. Metabolomics provides a comprehensive foundation for subtyping and precision psychiatry. Lipoxygenase-15, lipotoxicity, phospholipase A_2_, and lipid-redox intersections are important drug targets to treat MDD.

## Introduction

Major depressive disorder (MDD) is progressively regarded as a neuro-immune, oxidative, and nitrosative stress (NIMETOX) disorder, wherein peripheral immune-redox pathways induce metabolic alterations that result in neurotoxicity and modifications in limbic-prefrontal affective circuits (Maes, Almulla et al. 2025, Maes, Almulla et al. 2025, Maes, Jirakran et al. 2025). Metabolic anomalies are pivotal in the NIMETOX framework, as immune activation and oxidative/nitrosative stress (O&NS) alter lipid and energy metabolism, resulting in neurotoxic consequences rather than merely being secondary phenomena of depression (Maes, Galecki et al. 2011, Leonard and Maes 2012, Somani, Singh et al. 2022, Maes, Almulla et al. 2025). Oxygen and nitrogen species (ROS/RNS) and consequent O&NS impair lipids, proteins, and mitochondria, resulting in lipid peroxidation products, carbonyls, and redox-active metabolites that both indicate and exacerbate immune–oxidative toxicity (Maes, Galecki et al. 2011, Vaváková, Ďuračková et al. 2015). Pro-inflammatory cytokines and acute-phase mediators stimulate an excess of eicosanoids and oxidized phospholipids, resulting in a shift of membranes towards adrenic enrichment, which may be identified by metabolomics as lipid-remodeling signatures (Maes, Galecki et al. 2011, Morris and Maes 2014).

Recent studies demonstrate that MDD is linked to extensive modifications in amino acid, energy, lipid, and redox metabolism, encompassing branched-chain and aromatic amino acids, one-carbon pathways, carnitines, sphingolipids, and phospholipids (Leroy, Mac Donald et al. 2022). Initial untargeted plasma investigations revealed distinctive panels associated with alterations in lipids, tocopherols, immune metabolites, dihydroxyphenylacetic acid, and amino-acids (Kaddurah-Daouk, Boyle et al. 2011, Zheng, Chen et al. 2013, Liu, Yieh et al. 2016). Subsequent research indicated disruptions in lipid metabolic pathways, fatty acids, proinflammatory phospholipids, lysophosphatidylcholines, triglycerides, purinergic metabolites, choline, and neurosteroids (Chan, Suridjan et al. 2018, Kim, Lee et al. 2018, Cai, Cao et al. 2019). Metabolomics analysis may thus yield a quantitative assessment of NIMETOX activity, including upstream factors (inflammation and O&NS) into downstream lipid-redox phenotypes pertinent to the MDD phenome. In this perspective, metabolic anomalies may serve as both biomarkers and effectors. Furthermore, due to the variability in NIMETOX disturbances among patients, metabolomic signatures may facilitate biological subtyping (“metabotypes”), which is essential for precision psychiatry (Maes 2022).

We have recently concluded that the phenome of depression, comprising various subdomains including affective, anxiety, melancholia, chronic fatigue, fibromyalgia, autonomic, and vegetative symptoms, as well as illness recurrence and both current and lifetime suicidal behaviors, should be evaluated using newly developed machine learning-based phenome scores (Niu, Zhang et al. 2025). These include: a) a generalized factor derived from the symptom domain of the current index episode, termed overall severity of depression (OSOD), b) a singular group factor encompassing all psychosomatic domains (designated as psychosomatic symptoms), c) current suicidal behaviors, and d) the recurrence of illness index (ROI), which is predicated on the lifetime number of episodes and suicidal behaviors (Niu, Zhang et al. 2025). Nevertheless, no research has characterized the metabotypes of MDD and its associated features, including OSOD, physiosomatic symptoms, and suicidal ideation. Thorough metabolomics in meticulously characterized MDD cohorts is essential to discern distinct metabolic profiles that facilitate the influence of immune activation and redox mechanisms on neuro-affective toxicity.

Hence, this study aimed to a) identify metabolomic abnormalities distinguishing patients with MDD from healthy controls through high-resolution, untargeted mass spectrometry; b) enhance precision psychiatry by delineating biologically significant MDD metabotype signatures and metabolic fingerprints of MDD patients; c) examine associations between metabolomic alterations and clinical phenotypes, including OSOD, physiosomatic symptoms, current suicidal ideation, and ROI.

## Methods

### Participants

This study recruited 165 people, comprising 125 MDD inpatients and 40 healthy controls (HCs). This research employed a case-control cross-sectional design conducted at the International NIMETOX Center, Mental Health Center of Sichuan Provincial People’s Hospital in Chengdu, China. The participants’ ages ranged from 18 to 65 years, demonstrating an equitable gender ratio. Inpatients with MDD were diagnosed according to DSM-5 criteria, were in an acute phase of illness, and demonstrated a Hamilton Depression Rating Scale 21 (HAMD-21) score greater than 18 (Niu et al., 2025). The control group consisted of hospital staff, their relatives, and friends of patients, matched to the case group based on age, gender, education, and body mass index (BMI).

Participants in the control group were excluded if they had a diagnosis of MDD, dysthymia, DSM-5 anxiety disorders, or a familial history of mood disorders, substance use disorders (except nicotine dependence), or suicide. Exclusion criteria for this study included: a) diagnosis of other major psychiatric disorders such as schizophrenia, eating disorders, substance use disorders (except nicotine dependence), bipolar disorder, psycho-organic disorders, schizoaffective disorder, and autism spectrum disorder; b) neurological diseases such as epilepsy, stroke, multiple sclerosis, Alzheimer’s and Parkinson’s disease, and brain tumors; c) severe allergic reactions within the past month; d) experiencing an infection during the last three months; e) systemic conditions such as rheumatoid arthritis, inflammatory bowel disease, type 1 diabetes, chronic obstructive pulmonary disease, systemic lupus erythematosus, psoriasis, or cancer; f) present administration of immunosuppressants, corticosteroids, or other agents that modify the immune system; g) being pregnant or breastfeeding; h) surgery conducted within the past three months; i) antisocial or borderline personality disorder and developmental disorders; j) frequent consumption of analgesics; and k) use of therapeutic amount of antioxidants or Omega-3 supplements in the past three months; All participants or their legal representatives furnished written informed consent. The Ethics Committee of Sichuan Provincial People’s Hospital sanctioned the study [Ethics (Research) 2024-203].

### Clinical Assessment

A qualified physician conducted a semi-structured interview to gather demographic data, medical history, psychological history, and familial history. The validation of psychiatric diagnoses was performed using the Mini International Neuropsychiatric Interview (M.I.N.I.), which assesses a range of psychiatric lifetime and current disorders. On the same day, the assessor distributed a range of questionnaires to all participants to assess different rating scales measuring the intensity of somatic-psychosomatic symptoms, anxiety, and depression, as previously reported (Niu, Zhang et al. 2025).

The severity of MDD was assessed using the HAMD-21; anxiety severity was measured with the Hamilton Anxiety Scale; self-reported depression was quantified using the Beck Depression Inventory; and state anxiety was evaluated through the State-Trait Anxiety Inventory state subscale (Hamilton 1959, Hamilton 1960, Spielberger, Gorsuch et al. 1983, Beck, Steer et al. 1996). The Somatic Symptom Scale-8 (SSS-8) assessed the intensity of somatic symptoms experienced by individuals over the preceding seven days (Gierk, Kohlmann et al. 2014). The FibroFatigue Scale (FFS), a 12-item clinical interview tool, was utilized to assess the severity of symptoms related to chronic fatigue syndrome (CFS) (Zachrisson, Regland et al. 2002, Gierk, Kohlmann et al. 2014). The Columbia Suicide Severity Rating Scale (C-SSRS) assessed both lifetime and present suicidal attempts (SA) and suicidal ideation (SI) (Posner, Brown et al. 2011). These scales were used to compute OSOD, physiosomatic symptoms and current suicidal ideation scores, as explained previously (Niu, Zhang et al. 2025). This study utilized two items from the C-SSRS (number of lifetime suicidal attempts and ideation before the index episode) to compute the reoccurrence of illness (ROI) index. The ROI is a z unit-based composite score that is produced by adding the z-scores for the number of depressive episodes, suicidal attempts and suicidal ideation across one’s lifetime (Maes, Jirakran et al. 2024).

The current study analyzed the elements of metabolic syndrome (MetS), encompassing height, weight, body mass index (BMI), and waist circumference (WC). BMI was calculated by dividing weight in kilos by the square of height in meters. Waist circumference was measured midway between the iliac crest and the inferior rib. MetS was defined according to the 2009 joint statement from the American Heart Association and the National Heart, Lung, and Blood Institute (Alberti, Eckel et al. 2009). MetS was diagnosed when three or more of the following five criteria were met: (a) male waist circumference ≥90 cm or female waist circumference ≥80 cm; (b) triglycerides ≥150 mg/dL; (c) male HDL cholesterol <40 mg/dL or female HDL cholesterol <50 mg/dL; (d) systolic blood pressure ≥130 mm Hg or diastolic blood pressure ≥85 mm Hg, or use of antihypertensive medications; (e) fasting glucose ≥100 mg/dL or diagnosed diabetes mellitus.

### Assays

Blood samples were obtained between 06:30 and 08:00 hours, with 30 mL of fasting venous blood extracted using a reusable syringe and subsequently transferred into serum tubes. Following centrifugation at 3500 rpm, the serum was collected and aliquoted into Eppendorf tubes and stored at −80°C for later analysis. Samples were thawed on ice and vortexed for 10 s. 50 μL of sample and 300 μL of extraction solution (ACN: Methanol = 1:4, V/V) containing internal standards were added into a 2 mL microcentrifuge tube. The samples were vortexed for 3 min and then centrifuged at 12000 rpm for 10 min (4 °C). 200 μL of the supernatant was collected and placed in −20 °C for 30 min and then centrifuged at 12000 rpm for 3 min (4 °C). A 180 μL aliquots of supernatant were transferred for LC-MS analysis. Quality control (QC) was obtained by sucking 50μL of the supernatant of each biological sample and vortexing it in a centrifuge tube at room temperature for 60 seconds. Data acquisition and analysis of the experiment were performed at Beijing Junfeix Technology Co., Ltd. The LC-MS analyses were performed using a UPLC system (Vanquish, Thermo Scientific) coupled to an electrospray ionization quadrupole-Orbitrap hybrid high-resolution mass spectrometer (Q Exactive HF-X, Thermo Scientific).

All samples were examined using liquid chromatography (LC) and mass spectrometry (MS) methods. The LC-MS analyses were performed using a UPLC system (Vanquish, Thermo Scientific) coupled to an electrospray ionization quadrupole-Orbitrap hybrid high-resolution mass spectrometer (Q Exactive HF-X, Thermo Scientific). One aliquot was analyzed using positive ion conditions and was eluted from T3 column (Waters ACQUITY Premier HSS T3 Column 1.8 µm, 2.1 mm * 100 mm) using 0.1 % formic acid in water as solvent A and 0.1 % formic acid in acetonitrile as solvent B in the following gradient: 5 to 20 % in 1 min, increased to 99 % in the following 2 mins and held for 1.5 min, then come back to 5 % mobile phase B within 0.1 min, held for 1.4 min. The analytical conditions were as follows, column temperature: 40 °C; flow rate, 0.4 mL/min; injection volume: 4 μL. Another aliquot used negative ion conditions and was the same as the elution gradient of positive mode. All the methods alternated between full scan MS and data dependent MS-n scans using dynamic exclusion. MS analyses were carried out using electrospray ionization in the positive ion mode and negative ion mode using full scan analysis over m/z 75-1000 at 35000 resolutions. Additional MS settings are: ion spray voltage, 3.5 KV or 3.2 KV in positive or negative modes, respectively; Sheath gas (Arb), 30; Aux gas, 5; Ion transfer tube temperature, 320 °C; Vaporizer temperature, 300 °C; Collision energy, 30,40,50 V; Signal Intensity Threshold, 1.00E+06 cps; Top N vs Top speed, 10; Exclusion duration, 3s.

The acquired MS data pretreatments including peak picking, peak grouping, retention time correction, second peak grouping, and annotation of isotopes and adducts were performed using XCMS software. LC−MS raw data files were converted into mzXML format and then processed by the XCMS, CAMERA and metaX toolbox implemented with the R software. Each ion was identified by combining retention time (RT) and m/z data. Intensities of each peak were recorded and a three-dimensional matrix containing arbitrarily assigned peak indices (retention time-m/z pairs), sample names (observations) and ion intensity information (variables) was generated. The online KEGG, HMDB database was used to annotate the metabolites by matching the exact molecular mass data (m/z) of samples with those from database. If a mass difference between observed and the database value was less than 10 ppm, the metabolite would be annotated, and the molecular formula of metabolites would further be identified and validated by the isotopic distribution measurements. We also used an in-house fragment spectrum library of metabolites to validate metabolite identification.

## Statistics

### Classical statistical tests

Contingency table analysis was employed to investigate relationships among variables using categorical data, utilizing Chi-square testing as the methodological approach. An analysis of variance was utilized to assess the differences across the various study groups for continuous variables, including demographic and clinical data.

The sample size estimate was derived from the findings of Maes et al. (different papers), which indicated that a significant portion of the variance in OSOD, i.e., more than 25% could be anticipated using multivariable regression analysis including metabolic or NIMETOX variables. Power calculation for the primary statistical analysis of this study, specifically multiple regression analyses examining the predictors of the phenome scores, was conducted using G*Power 3.1.9.4 software. The analysis utilized an effect size of 0.33 (approximately 25% of the phenome explained), an alpha level of 0.05 (two-tailed), a power of 0.8, and a maximum of seven covariates. The power analysis indicated a minimum necessary sample size of 51 based on the determined effect size. The sample size was increased to include testing and holdout samples.

### Machine learning for biomarker discovery

Biomarker discovery was conducted utilizing a supervised multistage pipeline that incorporated principal component analysis (PCA), correspondence PCA, linear discriminant analysis (LDA), partial least squares discriminant analysis (PLS-DA), PLS-regression, neural networks (NN), support vector machines (SVM), and stringent sample splitting techniques to prevent model overfitting or circularity. Statistical analysis was predominantly performed utilizing R (v4.0), Statistica 14.0, SPSS 30.0, and the Unscrambler. Cluster heatmaps were produced using the R package heatmap. The volcano map and heatmaps were generated using the OmicStudio tool at https://www.omicstudio.cn/tool.

Metabolite data underwent three principal processing steps: first, data filtration to exclude samples with over 80% missing values or quality control (QC) samples with over 50% missing data; second, data imputation employing the K-nearest neighbor (KNN) method; and third, data standardization through Probabilistic Quotient Normalization (PQN). PCA on significant differential metabolite analysis was conducted with the R package metaX and the Unscrambler 14.0. PLS-DA was conducted using a) the R package ropls, which was followed by the calculation of variable importance in projection (VIP) values for each variable; and b) Statistica 14.0, which was followed by the computation of variable importances and case-wise score contributions.

CLR modifications were employed due to the compositional nature of metabolomics data, which is bound by constant-sum scaling, making it vulnerable to spurious correlations and misleading patterns when evaluated using raw or non-ratio-based scales. CLR eliminates compositional dependencies by representing each metabolite with relation to the geometric mean of all metabolites, hence generating data appropriate for linear multivariate techniques, and enhancing the interpretability of the covariance structure.

Analyses were conducted on the comprehensive, curated, and selected CLR-transformed datasets to investigate multivariate differentiation between MDD inpatients and healthy controls utilizing all or curated metabolites. Initially, we provide data from the comprehensive metabolomics matrix, which includes both endogenous and external metabolomics (n=736). Secondly, investigations were performed on the comprehensive curated metabolomics matrix comprising solely endogenous metabolites. Thirdly, selected metabolites were utilized in statistical studies (see below). The performance of the PLS-DA model in the metabolomic matrices of 736 and 112 metabolites was examined using R²Y (explained variance in group membership), Q² (cross-validated predictive performance), and cross-validated ANOVA (CV-ANOVA), which evaluated the statistical significance of class separation. To ascertain the variables most significantly influencing this multivariate structure, variable importance in projection (VIP) scores were obtained, and the study was repeated utilizing the highest-ranked VIP features to illustrate their combined impact on the PLS-DA discrimination. We will show results on PLS-DA applied to the complete data set (exogenous + endogenous) and VIP-selected metabolites from the curated data set. This approach was performed exclusively for exploratory multivariate analysis, not for biomarker identification, predictive modeling, or validation.

During cross-validation methods, all feature selection, model training, hyperparameter optimization, and interpretability processes were conducted solely within the training set, with the testing and holdout samples designated entirely for final assessment. This design mitigated leakage at every analytical tier, guaranteeing impartial biomarker identification and reliable predictive modeling (Kuhn and Johnson 2013). The complete sample was randomly divided into training (50%), testing (25%), and holdout (25%) sets. The preliminary biomarker screening was confined to the training sample. All data inside the training samples underwent scaling by Pareto or z transformations. The data were further cross-validated in the testing sample with PLS-DA, SVM, and neural networks, which included a holdout sample. No VIP-selected metabolite was utilized in the development of classification models, thus circumventing leakage and ensuring complete independence between exploratory multivariate modeling and subsequent biomarker analysis.

In the training subset, each metabolite was assessed using t-tests or univariate ANOVAs to identify differences between MDD and control groups, with adjustments made for potential confounders such as age, sex, BMI, and metabolic syndrome. The final notable differential metabolites were identified according to three criteria: P-value < 0.05 from t-tests or ANOVAs, fold change >1.2, and VIP ≥1 from PLS-DA analysis. Consequently, 46 metabolites were obtained. The candidate list was then diminished through repeated refinement to ten metabolites. Subsequently, an automated stepwise LDA was executed on the training sample to ascertain six variables that uniquely contributed to class discrimination above and beyond the 10 delineated metabolites. The amalgamation of ANOVA-significant features and LDA-selected features yielded a compilation of 16 potential metabolites. Utilizing these 16 metabolites, we developed six z-based composite scores that represent six distinct functional metabolic profiles.

### Testing procedures

The 16 primary variable scores and 6 derived domain scores were subsequently utilized in the real model evaluation phase inside the testing population. To evaluate the replicability of categorization, we initially conducted a z transformation of the data in the testing set and recalculated the composite scores within this restricted sample. The trained models, utilizing either 16 or 6 variables, were subsequently subjected to LDAs applied solely to the testing sample, employing leave-one-out (LOO) cross-validation to assess out-of-sample prediction stability. A feed-forward neural network was trained on the training set and assessed on the independent testing set, with performance also analyzed in the holdout sample, which had not been utilized in any phase of training, feature selection, or tuning. A support vector machine (SVM) classifier was employed utilizing a 10-fold cross-validation methodology to assess the resilience and generalizability of predicted accuracy over various resampling folds. Only performance estimates obtained from the amalgamated testing and holdout samples or from cross-validated SVM models were deemed valid indications of predictive utility. The technique maintained a rigorous separation between variable selection and model evaluation, so assuring unbiased biomarker identification and preventing data leaking.

### Regression methods

We utilized PLS-regression to analyze the prediction of phenome scores (OSOD, present SI, physiosomatic symptoms) in the aggregated training and testing sets. The model’s accuracy was assessed using R²Y alongside Q² values for all obtained latent vectors. Thus, we calculated score contribution profiles or case-levels contributions for each participant. This method allows visualization of how specific metabolites push a given participant toward increased phenome scores. This provides a mechanistic rationale for each prediction and allows identification of which specific metabolic abnormalities drive severity of illness in each case, an approach that is increasingly recognized as essential for personalized precision psychiatry.

Alongside the manual multiple regression method, we employed automated regression techniques to ascertain the metabolic scores that operate as predictors for phenome attributes. This inquiry conducted a ridge regression analysis using a regularization parameter of λ=0.1 and a tolerance level of 0.4, using Statistica, Windows version 14. Additionally, we employed forward stepwise automatic linear modeling analyses, implementing an overfit criterion for variable inclusion and exclusion, with a maximum effects threshold established at 5 (using SPSS, Windows version 30). We performed a “best subset” analysis using a criterion to prevent overfitting, focusing on the 8 most significant metabolites revealed in the aforementioned regressions conducted with SPSS version 30. Subsequent to these assessments, we executed a manual regression analysis utilizing SPSS 30 and ran an exhaustive examination of the model statistics, encompassing the F statistic, degrees of freedom, and p-values, in addition to the total variance elucidated by R². Additionally, we conducted a thorough study of the standardized beta coefficients for each predictor, along with their corresponding t statistics and exact p-values. The final models were assessed for multivariate normality, encompassing the residual distributions and P-P plots. An assessment of the variance inflation factor and tolerance was conducted to identify any potential issues related to collinearity or multicollinearity. The evaluation of heteroskedasticity was performed using the White test and the modified Breusch-Pagan test to determine homoscedasticity. Additionally, we performed partial regression analyses of phenome data related to metabolites. The specified regression analyses were performed using IBM SPSS, Windows version 30 and Statistica, version 14.0. The significance threshold for all statistical analyses was established at 0.05, employing two-tailed tests.

## Results

### Demographic and clinical data

Electronic Supplementary File (ESF) Table 1 shows that there were no significant differences in age, sex distribution, BMI, MetS prevalence, and education between MDD inpatients and controls. MDD patients showed increased OSOD, physiosomatic, suicidal ideation and ROI scores as compared with controls.

**Table 1.** The top 16 selected metabolites and the 6 derived metabolic functional modules.

| Index used in the current study | Annotations | Sign in depression | Functional group |
| --- | --- | --- | --- |
| ME0050035 | 1-Stearoyl-2-arachidonoylglycerol | Increased | Lipotoxicity |
| ME0050075 | DG(18:1(11Z)/18:2(9Z,12Z)/0:0) | Decreased | Lipotoxicity |
| ME0055180 | Myristic acid, tetradecanoic acid, C14:0 | Increased | Lipotoxicity |
| ME0168773 | Arachidic acid, eicosanoic acid, C20:0 | Decreased | Changes in FA storage/<br>metabolism/signaling |
| ME0054914 | Methyl stearate, methyl C18:0 | Increased | Changes in FA storage/<br>metabolism/signaling |
| ME0054522 | 1-O-Hexadecyl-lyso-sn-glycero-3-phosphocholine, Lyso PAF-C-16 | Decreased | Ether lipids affected by<br>immune-redox stress |
| ME0061811 | PI(22:4(7Z,10Z,13Z,16Z)/16:0), PI 16:0/22:4 | Increased | PL remodeling |
| ME0061712 | 14,15-Leukotriene C4(ExC4), Eoxin C4 | Increased | PL remodeling |
| ME0063012 | PS(22:5(4Z,7Z,10Z,13Z,16Z)/22:4(7Z,10Z,13Z,16Z)) | Increased | PL remodeling |
| ME0012993 | 1-Stearoyl-2-arachidonyl-sn-glycero-3-phosphocholine | Increased | PL remodeling |
| ME0063041 | PS(22:5(7Z,10Z,13Z,16Z,19Z)/22:4(7Z,10Z,13Z,16Z)): | Increased | PL remodeling |
| ME0141123 | 1,2-Dioleoyl-sn-glycero-3-phospho-(1'-myo-inositol) | Increased | PL remodeling |
| ME0063301 | rac-4-Hydroxy-4-O-(beta-D-glucuronide)-all-trans-retinyl acetate | Decreased | Retinoid detox, Vit A<br>metabolism |
| ME0148101 | Cysteylhistidine | Increased | Mito ATP Redox |
| ME0103600 | Inosinic acid | Increased | Mito ATP Redox |
| ME0118596 | Hydroxyphenyllactic acid | Increased | Mito ATP Redox |

FA: fatty acid

### Discrimination of MDD versus controls

The initial metabolomics dataset had 736 identified features. ESF, Figure 1 shows the volcano plot, utilizing log2 fold change, of 736 metabolites reveals a significant pattern of differential regulation in MDD. Fifty-nine metabolites were considerably elevated, whereas one hundred fifty-five metabolites were significantly reduced. The residual metabolites exhibited no notable variation, establishing a neutral backdrop against which the altered species are contrasted. PLS-DA conducted on the complete dataset of 736 metabolites demonstrated a significant multivariate distinction between MDD patients and control participants (refer to Electronic Supplementary File, Figures 2). The PLS-DA model demonstrated adequate performance metrics with Q² = 0.84, signifying robust predictive capability, R²Y = 0.918, indicating considerable explained variance in group classification, and R²X = 0.115, implying that a modest fraction of total metabolic variance is adequate for distinguishing the two groups. Permutation testing validated that the reported model performance was not attributable to random structure, as all permuted Q² and R² values were significantly worse to the empirical model.

**Figure 1.**
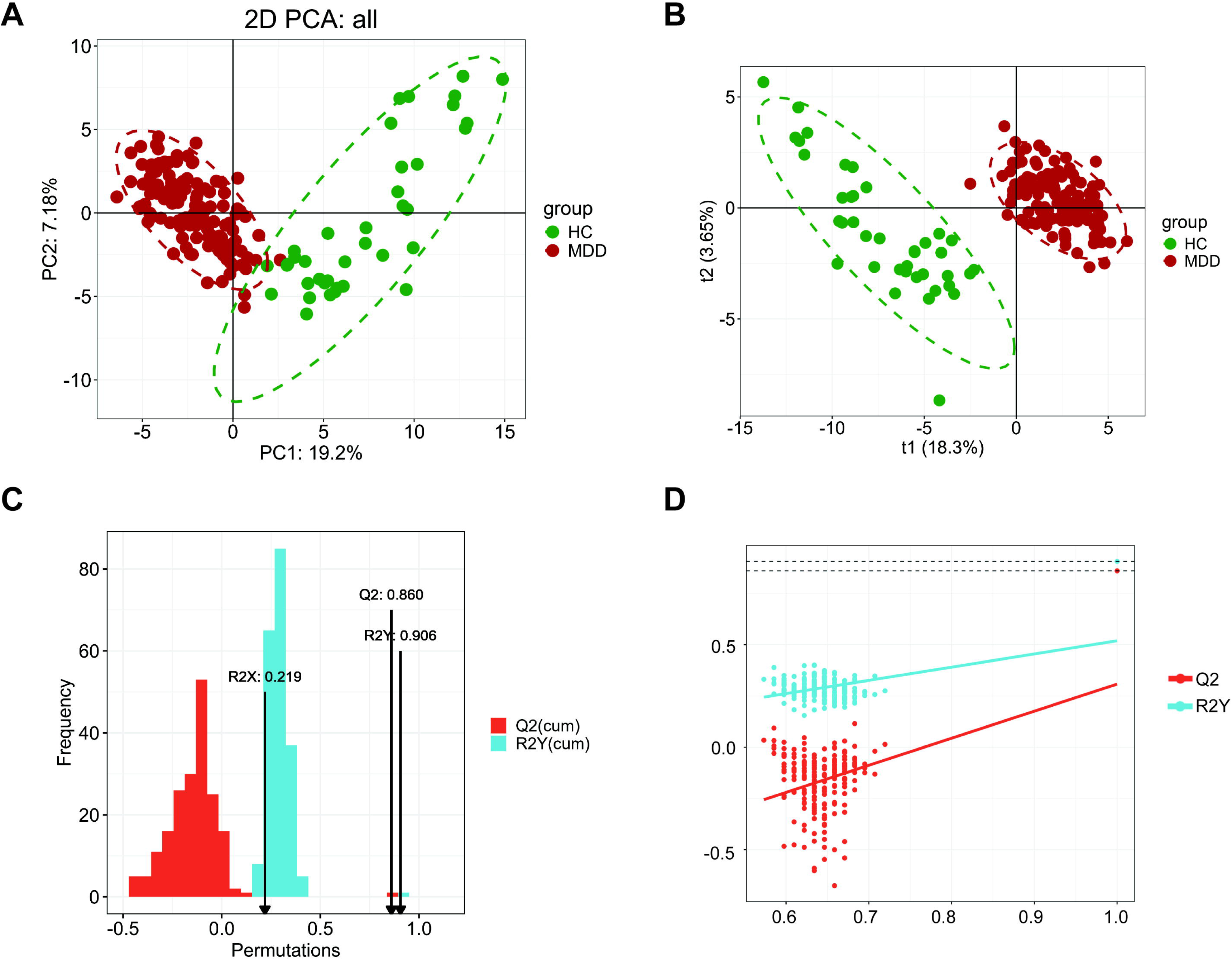
Results of principal component (PC) analysis and partial least squares discriminant analysis (PLS-DA). **Figure 1A** illustrates the principal component plot (PC). **Figure 1B** illustrates the PLS-DA model. **Figure 1C** shows the predictive performance with Q² and R²Y values, including after permutations. **Figure 1D** shows the permutation plot revealing the R^2^Y and Q² intercepts.

**Figure 2.**
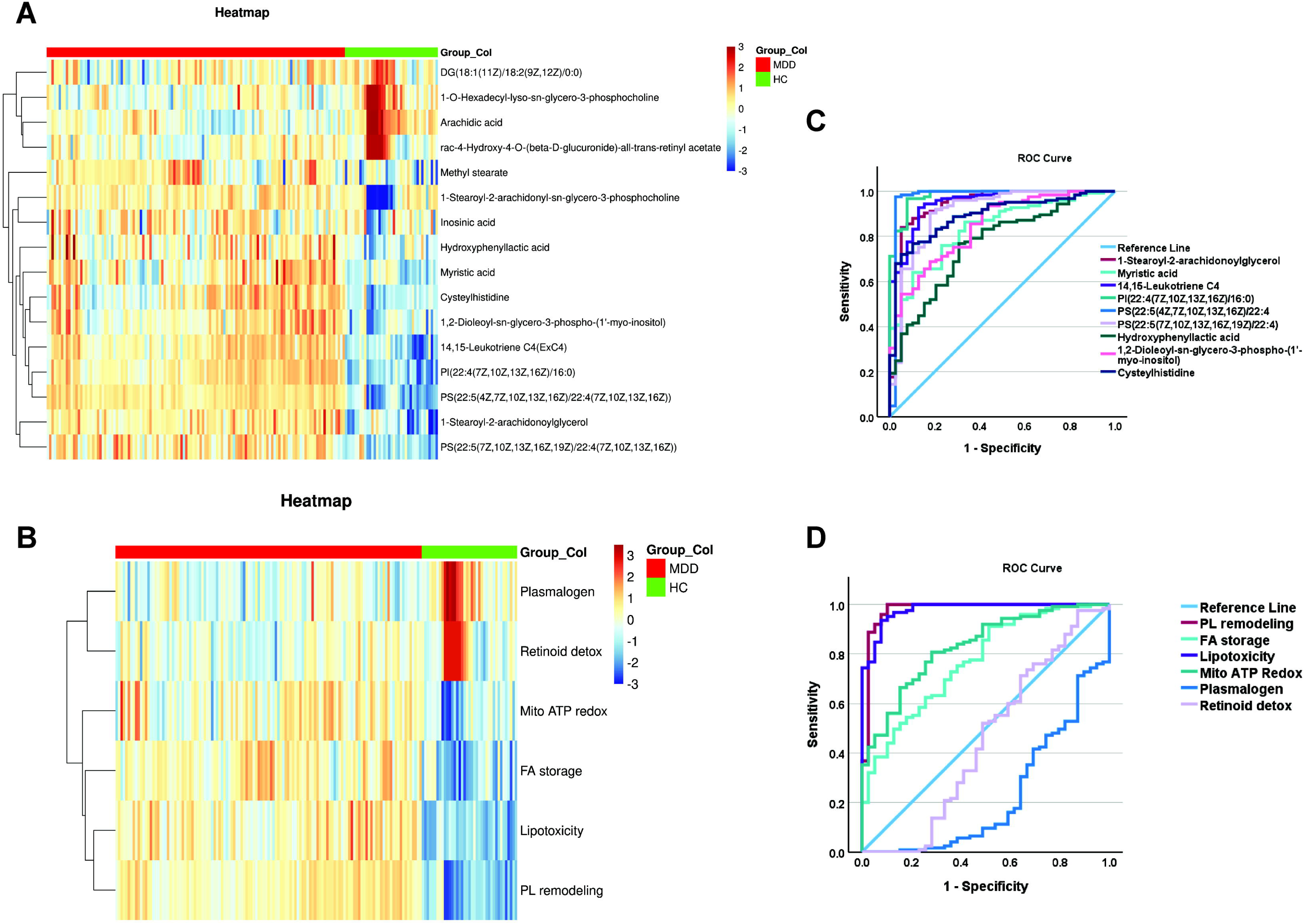
Heatmaps using the 16 top metabolites (**Figure 2A**) and the 6 derived functional modules (**Figure 2B**) as well as the receiving operating (ROC) curves showing the accuracy of the top metabolites (**Figure 2C**) and the functional domains (**Figure 2D**).

We excluded non-endogenous, exogenous, drug-related, artifact-derived, or low-confidence metabolites and preserved only those with VIP > 1.2 and FDR < 0.05. Examining the refined data set (n=112 high-confidence metabolites) showed that the multivariate structure remained entirely congruent with the PLS-DA analysis performed on 736 variables. PCA conducted on this modified dataset demonstrated a distinct and consistent distinction between MDD and control. **Figure 1A** illustrates that in the PC1 versus PC2 score plot, the two groups exhibit discrete, non-overlapping clusters. PC1 appeared as the principal discriminating axis. **Figure 1B** illustrates the PLS-DA model, which demonstrates two completely distinct clusters, with MDD and control patients residing in separate, non-overlapping areas of the score plot. The separation was characterized by a wide, vacant “street” between the clusters, signifying robust multivariate discrimination without transitional overlap. The modified model exhibits a significantly superior discriminating structure compared to the model presented in ESF, Figure 1, with t1 = 18.3% versus 8.69%, demonstrating that the elimination of non-endogenous elements and noise greatly improved the biological signal. Correspondingly, R²X rose from 0.115 to 0.218, indicating that a greater fraction of total metabolic variation is now encompassed by the latent components. The predictive performance (**Figure 1C**) was accurate, with Q² = 0.86, nearly identical to the original Q² = 0.84, and R²Y = 0.906, indicating significant class separation. The permutation plot (**Figure 1D**) reveals an intercept Q² significantly below zero, indicating that all permuted models underperformed relative to chance, hence demonstrating the absence of overfitting. The R²Y intercept of approximately 0.25 is significantly lower than the empirical R²Y, indicating that genuine class distinction cannot be attributed to random structure. Furthermore, CV-ANOVA exhibited highly substantial class differentiation, confirming the robustness and non-randomness of the PLS-DA model (p<0.0001). The ROC curve achieved a score of 100%. However, this ROC value cannot be construed as indicative of predictive performance, as it was computed from the combined training and testing dataset and, therefore, reflects resubstitution rather than true generalization. Accordingly, we do not use this ROC curve to claim diagnostic accuracy.

### Selection of the top metabolites and functional metabolic domains

As mentioned in the Statistics section, an initial training-testing split was established, with a distinct holdout sample allocated for final validation in neural networks. We performed univariate ANOVA on the CLR-transformed and scaled metabolomics data within the training dataset to find compounds with the most significant group discrimination. This technique identified the ten most discriminating variables based exclusively on statistically significant differences between MDD and control participants in the training sample. Concurrently, we conducted PLS-DA to elucidate multivariate structure and employed PLS regression with OSOD as the dependent variable. The multivariate models discovered six additional metabolites that significantly contributed to class separation and clinical severity, regardless of the features identified by ANOVA. The integration of these six multivariate predictors with the top 10 ANOVA-derived indicators resulted in significantly enhanced discrimination, indicating substantial additive effects among different metabolic pathways. The analysis yielded a final panel of 16 critical metabolites, as shown in **Table 1**, which represent the most useful biomarkers identified through both univariate and multivariate modeling in the training sample.

Utilizing the 16 discriminating metabolites, we developed six subdomain scores through standardized z-unit composite indices. **Table 1** delineates the six domains along with the assignment of specific metabolites to each, based on their recognized biochemical and pathological roles (see Discussion). The clusters represent cohesive processes: lipotoxicity, membrane phospholipid (PL) remodeling, fatty acid (FA) storage/signaling and redistribution, ether-lipid metabolism, mitochondrial-redox stress, and deficiencies in vitamin A and retinoid detoxification.

The heatmaps of the 16 selected metabolites and 6 functions in the training + testing samples demonstrate a distinct and consistent differentiation between MDD patients and controls (**Figure 2A and 2B**). MDD samples exhibit discrete, block-wise clusters of upregulated and downregulated metabolites across the matrix. The two groups display contrasting color gradients across many metabolic categories, signifying extensive yet organized metabolic reprogramming. This bidirectional, systematic difference validates a strong, group-specific metabolic signature.

### Predictive validity of the training-set model

PCA conducted on the testing sample utilizing the 16 metabolites demonstrated a distinct distinction between MDD and control groups within a two-dimensional space, accounting for 77% of the total variance (ESF, Figure 4). **Figure 2C and /2D** illustrate the ROC curves for the ten metabolites positively correlated with MDD and the six functional domains in the testing sample.

The ROC curves indicate AUC values surpassing 0.90 for some metabolites and functions, indicating robust univariate discrimination capability independent of multivariate analysis. The elevated AUCs validate the reliability of the chosen metabolites, independent of overfitted or non-cross-validated performance assessments.

The SVM analysis employing a kernel basis function, utilizing the 16 selected metabolites, attained a significant differentiation between MDD and HC. The kernel’s width value was 0.063, signifying a very adaptable decision boundary capable of representing nonlinear class structures. The final model employed 28 support vectors (SVs), 18 of which were bounded, indicating vectors positioned on or beyond the soft-margin boundary that actively influenced the classification hyperplane. The class composition of support vectors was equitable, comprising 15 from the MDD group and 13 from the control group, indicating a symmetrical contribution of both classes to the decision function. Ten-fold cross-validation produced an accuracy of 100%, demonstrating consistency in separation across all folds.

**Table 2** and **Figure 3** present the outcomes of neural networks using the 16 metabolites and 6 functional domains. The 16 variables (Figure 3A, Table 2, model #1) exhibited discriminating efficacy between MDD and healthy controls. The architecture comprised two hidden layers, featuring 6 units in the first layer and 5 units in the second, both employing the hyperbolic tangent activation function to examine nonlinear interactions among metabolites. The output layer comprised two units representing the two diagnostic categories (MDD versus control), with an identity activation function alongside a sum-of-squares error function. Model convergence was attained via the training-error ratio stopping criterion (0.001), signifying sustained optimization without indications of overfitting. The final error metrics were adequate, with a sum of squares of 0.011 in the training sample and 0.005 in the testing sample, indicating a good fit. The classification performance was consistently adequate, with an incorrect prediction rate of 0% across the three datasets. Figure 3A illustrates the significance chart, wherein Methyl stearate emerges as the predominant metabolite, succeeded at a considerable interval by increasing Hydroxyphenyllactic acid, followed distantly by 1-Stearoyl-2-arachidonoylglycerol, PS(22:5(4Z,7Z,10Z,13Z,16Z)/22:4(7Z,10Z,13Z,16Z)), and 14,15-Leukotriene C4 (eoxin C4).

**Figure 3.**
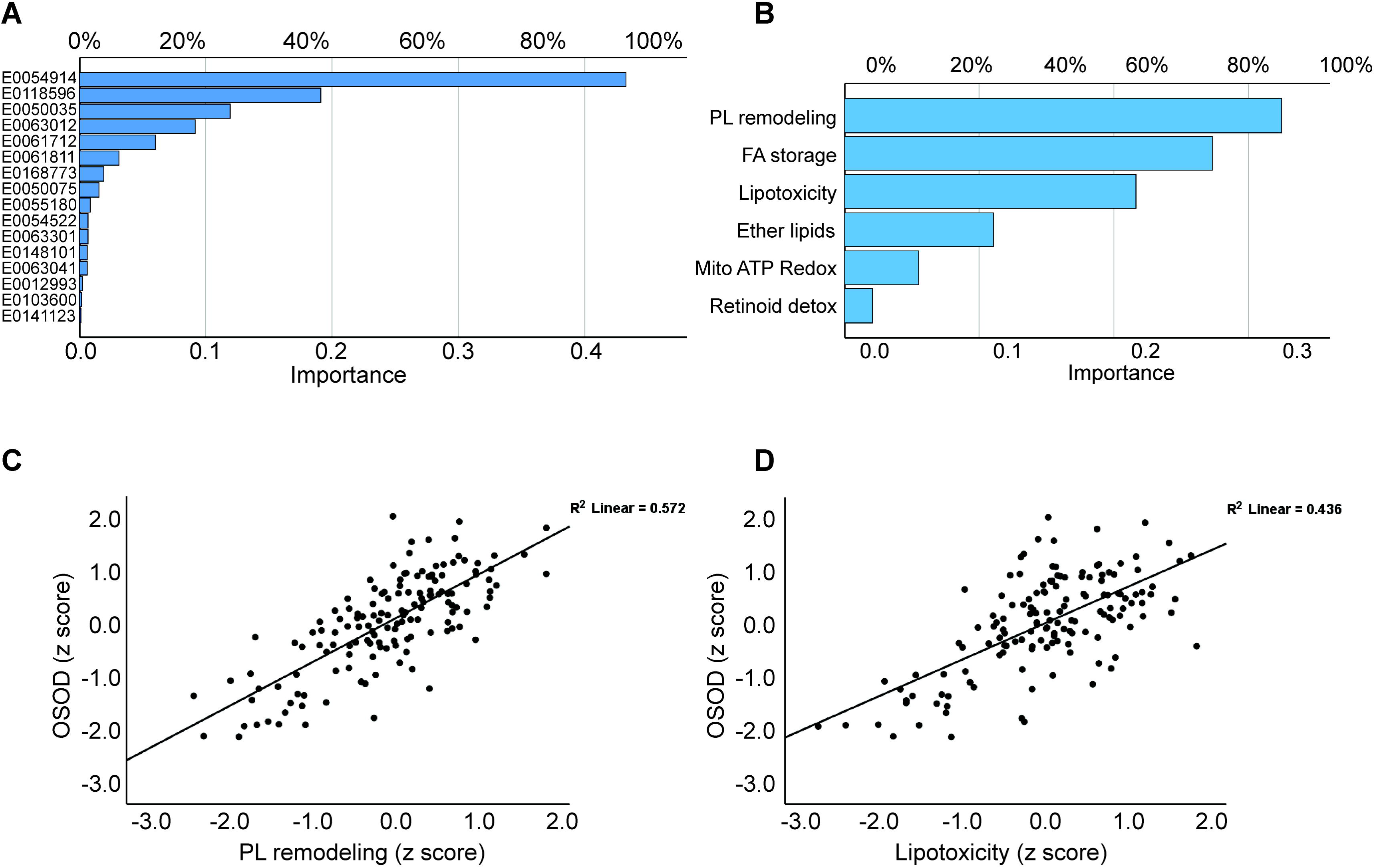
Results of neuronal network analysis showing the most relevant metabolites (**Figure 3A**) and the functional domains (**Figure 3B**). This figure shows the partial regressions of the overall severity of illness on phospholipid (PL) remodeling (**Figure 3C**) and lipotoxicity (**Figure 3D**).

**Table 2.**
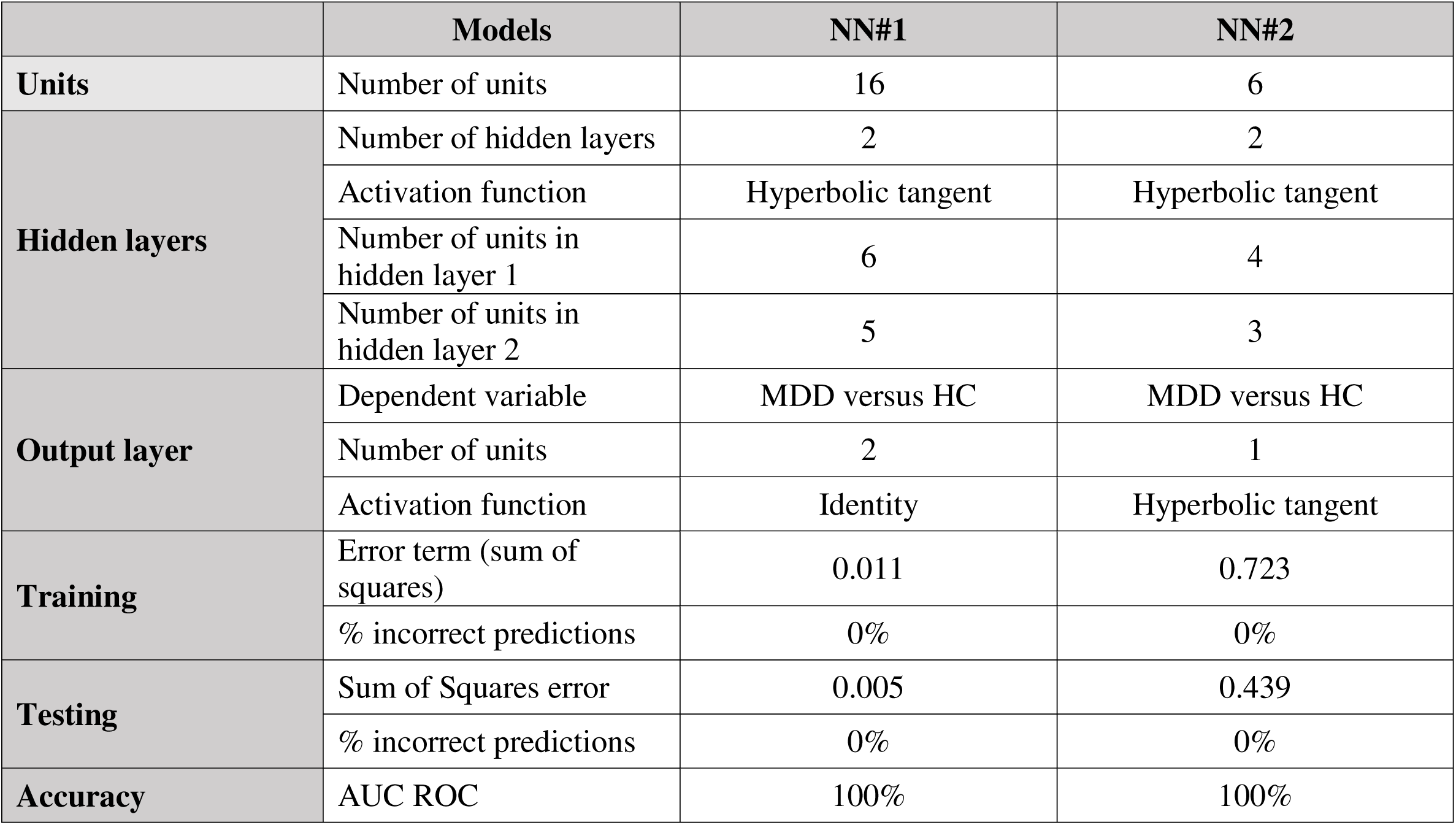
Results of neural networks (NN) with major depression (MDD) and healthy controls (HC) as output variables and either 16 selected metabolites or 6 metabolic functional domain scores as input data.

The neural network analysis of the six metabolomic functional domains (Figure 3B) demonstrated significant differentiation between MDD and controls. The network architecture had two hidden layers, featuring 4 units in the first layer and 3 units in the second, both employing hyperbolic tangent activation to represent nonlinear functional interactions. The output layer had two identity-activated units representing diagnostic classes, optimized by a sum-of-squares error function. Model convergence was achieved after one consecutive step without additional error reduction, signifying swift stabilization of the learning process. The resultant model had low sum-of-squares values (0.723 in training and 0.439 in testing), indicating adequate fit in both samples. The rate of erroneous predictions was 0% across the three samples. Figure 3B illustrates the significance chart, highlighting PL remodeling, FA storage/signaling, and lipotoxicity as the predominant functions, with ether-lid metabolism following at a distance.

LDA was utilized on the six metabolomic functional domains to assess their capacity to differentiate patients with MDD from healthy controls. As described above, composite scores for each functional domain in the training sample were initially created and then normalized by z-transformation to ensure scale comparability. Subsequently, LDA was conducted on these z composites in the training sample. This model exhibited significant group differentiation, evidenced by a Wilks’ lambda of 0.097 (χ² = 268.09, df = 6, p < 0.001) and a canonical correlation of 0.950, signifying that the discriminant axis accounted for the majority of variation between the MDD and control groups. The coefficients of the classification function obtained from this training model were subsequently applied to the independent testing sample. In the test set, metabolites were z scaled and composite scores were re-evaluated and z-transformed before utilizing the discriminant functions produced from the training data. The discriminant analysis in the testing sample demonstrated adequate separation, with Wilks’ lambda = 0.088 (χ² = 284.235, df = 6, p < 0.001) and a canonical correlation of 0.988. The predictive performance was maintained: total accuracy attained 97.5%, with a sensitivity of 95%, specificity of 100%, and a ROC AUC of 1.00. The results validate that the six functional metabolomic domains yield a highly consistent and distinctive signal for MDD across independent samples.

### Multiple regression analysis

**Table 3** displays the outcomes of several regression analyses, whereby OSOD, physiosomatic symptoms, suicidal ideation, and ROI serve as dependent variables, while the six lipidomic subdomains function as predictors. OSOD was predicted (72.5% of variation explained) by PL remodeling, FA storage/signaling, and lipotoxicity, all exhibiting positive correlations. **Figure 3C and 3D** illustrate the relevant partial regression plots for the regression of OSOD on PL remodeling and Lipotoxicity, respectively (adjusted for age, sex, MetS, and BMI). Model #2 indicates that physiosomatic symptoms were accounted for by 55.8% through the same three subdomains. Model #3 indicates that current suicidal ideation was predicted at 23.6% by PL remodeling and FA storage/signaling, both exhibiting positive regression coefficients. These findings underscore a common metabolic substrate that correlates with overall symptom severity, psychosomatic burden, and suicidal ideation.

**Table 3.**
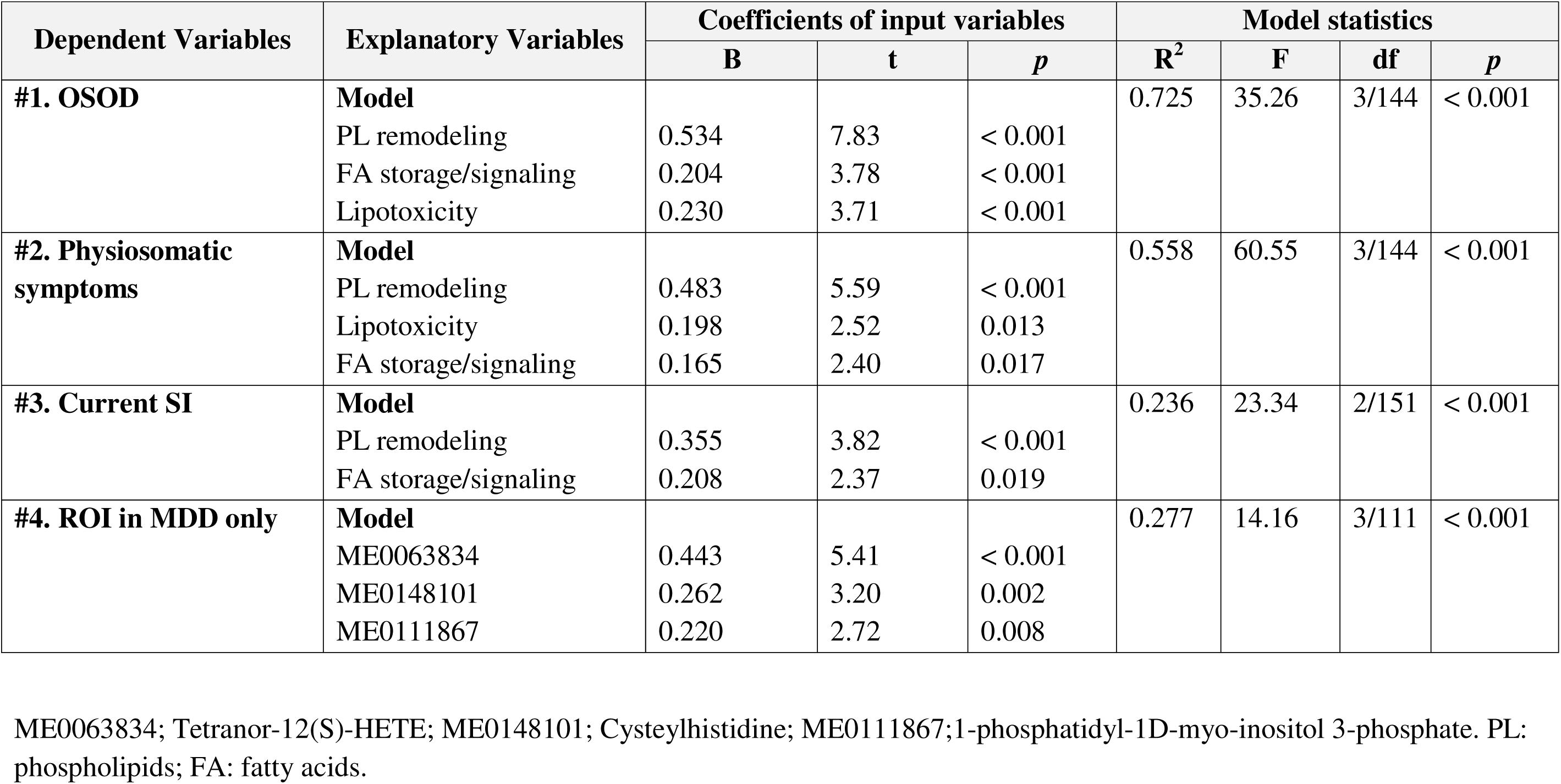
Results of multiple regression analysis with the overall severity of illness (OSOD), physiosomatic symptoms, current suicidal ideation (SI), and recurrence of illness (ROI) as dependent variables, and metabolites or functional module scores as explanatory variables.

Model #4 shows the regression of ROI on the metabolomics in the restricted group of MDD (to appreciate the effects of ROI in MDD). Only one of the 16 selected metabolites (i.e., ME0148101: Cysteylhistidine) was significant and, therefore, we examined whether other top metabolites could contribute. This model shows that 27.7% of the variance in ROI in MDD was associated with higher ME0063834, ME0148101, and ME0111867.

### Personalized approach via PLS-regression

A PLS regression was performed on the 16 selected metabolites within the total study cohort, utilizing OSOD as the dependent variable and the 16 principal metabolomic biomarkers as predictors. The model discerned two notable latent components, exhibiting eigenvalues of 6.54 and 1.33, respectively. The initial component explained a significant percentage of the variance in symptom severity (R²Y = 0.719), but the subsequent component accounted for a minor additional fraction (R²Y = 0.038). The model demonstrated predictive capacity, with Q² = 0.696 for the first component and a cumulative Q² = 0.707 with the addition of the second component. The cumulative explained variance in the predictors (R²X) was 0.413 and 0.509 for the two components, signifying that nearly half of the metabolomic variance significantly contributed to symptom prediction.

Figure 4 illustrates the variable importance metrics, indicating that several metabolites—including PI(22:4(7Z,10Z,13Z,16Z)/16:0), PS(22:5(4Z,7Z,10Z,13Z,16Z)/22:4(7Z,10Z,13Z,16Z)), and 14,15-Leukotriene C4 (eoxin C4)—exerted significant influence on OSOD. We calculated score contribution profiles for each participant to demonstrate the metabolite contributions. Figure 5 illustrates sample profiles from six individuals, comprising two controls and four MDD inpatients. Significant heterogeneity was noted among patients: some exhibited substantial positive contributions from E0148101 or E0141123, while others were defined by elevated E0050035, diminished E0168773, or unique combinations of these characteristics. A comparable pattern manifested throughout the six lipidomics functional domains (Figure 6), corroborating that, despite a common metabolomic signature of MDD at the group level, patients exhibit unique mechanistic subprofiles, indicating significant interindividual metabolic variability in disease manifestation.

**Figure 4.**
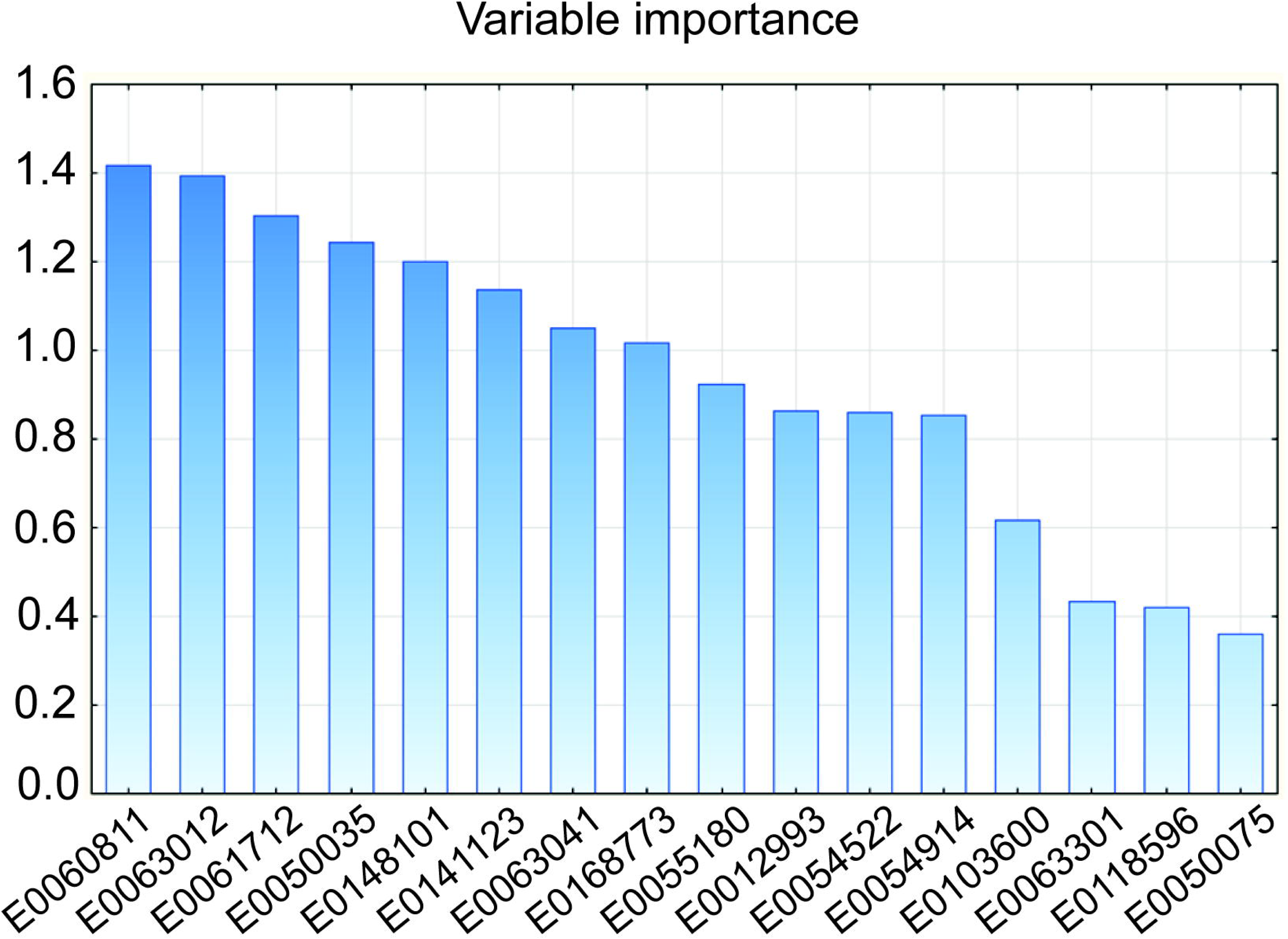
Results of partial least squares regression analysis with overall severity of depression (OSOD) as the dependent variable. This figure shows the importance of the 16 top metabolites for OSOD.

**Figure 5.**
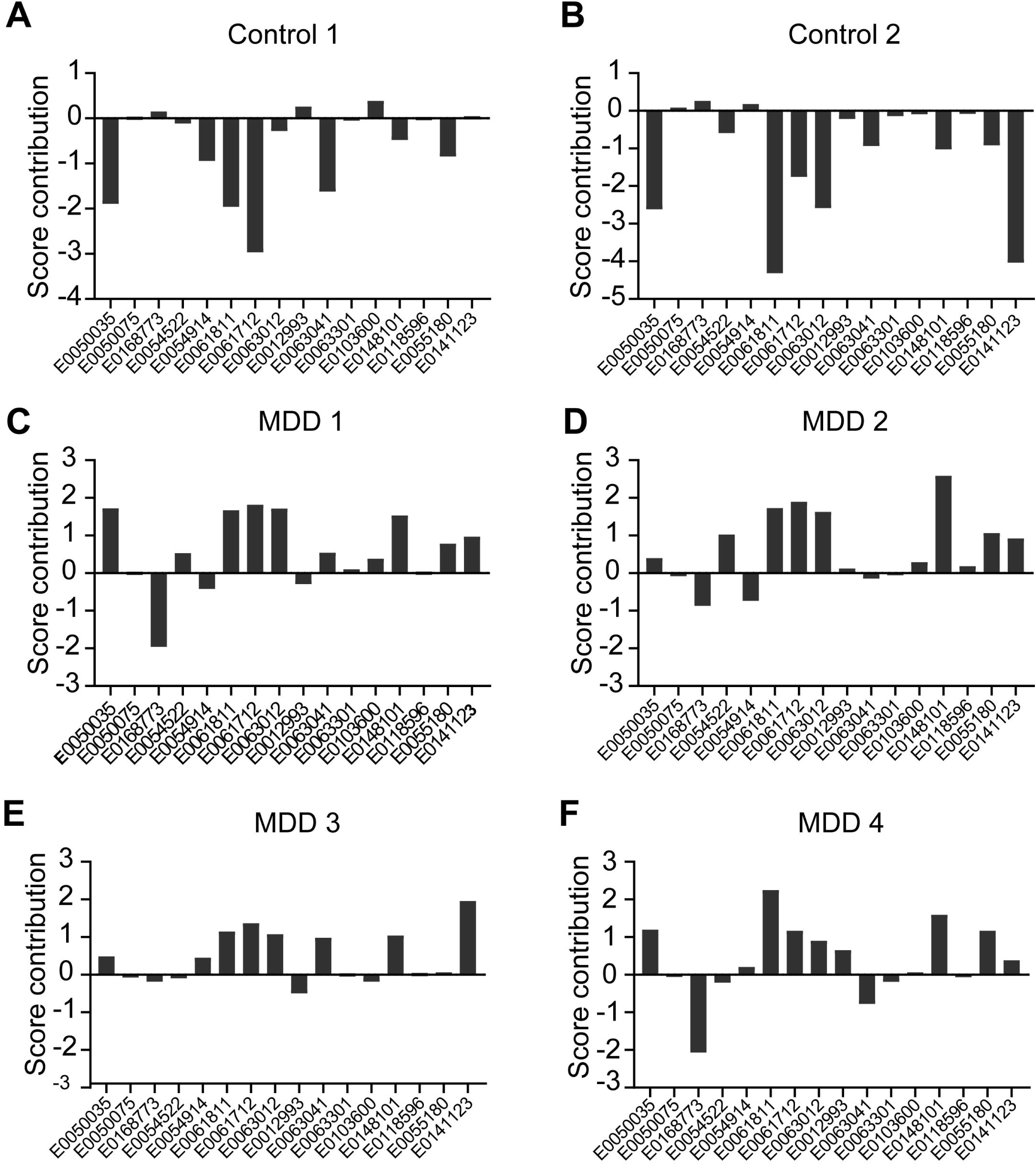
Results of case-wise score contributions based on the top 16 metabolites following partial least squares regression analysis with overall severity of depression (OSOD) as the dependent variable in six participants. Two healthy controls (**Figures 5A** and **5B**) and 4 inpatients with major depressive disorder (**Figures 5C, 5D, 5E** and **5F**).

**Figure 6.**
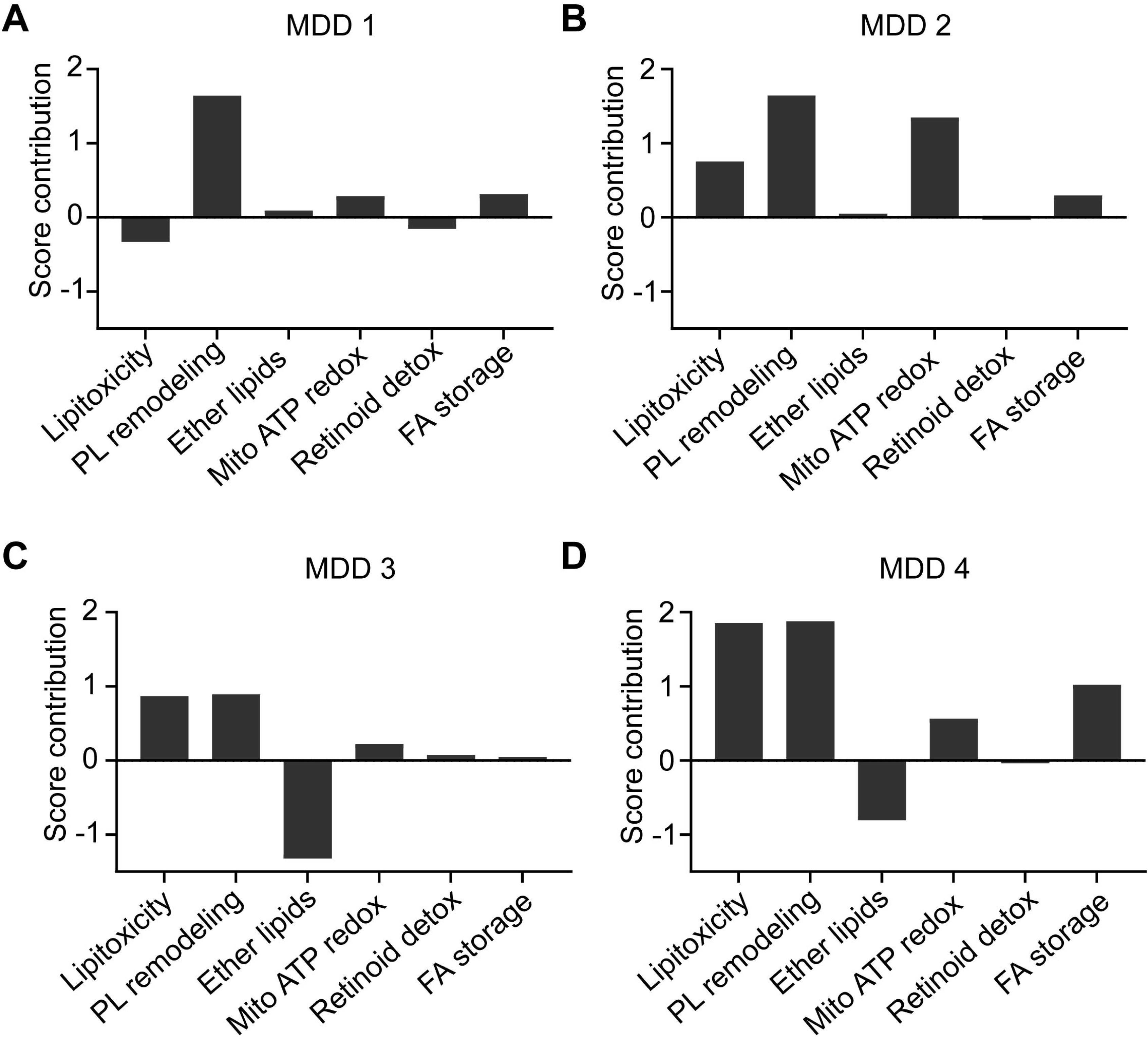
Results of case-wise score contributions based on the 6 functional domains following partial least squares regression analysis with overall severity of depression (OSOD) as the dependent variable in four inpatients with major depressive disorder (**Figures 6A, 6B, 6C** and **6D)**.

### Effects of confounding variables

Table 1 of the ESF indicates that the prevalence of MetS was comparable across individuals with MDD (17.7%) and controls (25.6%). MetS exhibited no significant impact on the 16 metabolites (F=0.92, df=116/142, p=0.553) or the 6 functional domains (F=1.23, df=6/152, p=0.295; results from multivariate GLM analysis). Moreover, MetS exhibited no impacts on any of the 16 metabolites or 6 domains, even in the absence of p-value correction. The regression of OSOD, physiosomatic symptoms, suicidal ideation and ROI on the metabolites (Table 3) remained unchanged after forced inclusion of MetS, BMI, age, and sex. In addition, the same results (16 metabolites and 6 functional modules) were detected in subjects without MetS. Our observations reveal that 92 patients with MDD were treated with antidepressants, 68 were prescribed benzodiazepines, 44 received atypical antipsychotics, and 10 were supplied with mood stabilizers. These medications did not produce any significant effect on the 16 metabolites or the 6 domains, even without FDR p-value correction. Consequently, there is no data indicating that the patient’s medication condition influenced the outcomes of the study.

## Discussion

### Distinct metabolic profiles underlying MDD

The primary finding of this work is that the obtained metabolomic signatures consistently and reliably represent a stable metabolic phenotype of MDD across several analytical dimensions. The comprehensive dataset, the condensed panel of 112 features, the 16 variables selected from the training sample, and the constructed 6 functional domains produced accurate models, affirming the existence of a metabolic signal linked to MDD and its phenome features. Furthermore, the metabolic features to a large extent predict OSOD, physiosomatic symptoms, and suicidal ideation during the index episode. The ROI index was strongly associated with a metabolic module that might further aggravate the NIMETOX pathways (Maes, Almulla et al. 2025). Therefore, the metabolomic profiles of MDD and its phenome features align with a consistent NIMETOX-associated metabolic phenotype, which will be elaborated upon in the subsequent section.

Prior systematic metabolomics or lipidomics investigations have demonstrated that MDD is associated with a multisystem metabolic abnormality. A preliminary NMR/MS investigation by Kaddurah-Daouk and colleagues revealed alterations in dihydroxyphenylacetic acid, tocopherols, and serotonin (Kaddurah-Daouk, Boyle et al. 2011). Subsequent LC–MS plasma analyses conducted by Chinese researchers identified modifications in lipids, immune-related metabolites, stress hormones, amino acids, and catecholamines, with panels distinguishing MDD from controls exhibiting an area under the curve (AUC) of around 0.8–0.9 (Zheng, Chen et al. 2013, Liu, Yieh et al. 2016). Further investigation uncovered modifications in fatty acids, purinergic metabolites, choline, and neurosteroids (Cai, Cao et al. 2019). Chan et al. discovered data indicating the involvement of proinflammatory phospholipid species in the occurrence of depression among coronary artery disease patients (Chan, Suridjan et al. 2018). Kim et al. (2018) discerned a lipid signature of MDD marked by alterations in lysophosphatidylcholines and triglycerides (Kim, Lee et al. 2018). Multi-omics profiling of animal models of MDD reveals significant disruptions in cerebellar energy metabolism, amino acid metabolism, heightened expression of glycolytic and TCA-cycle enzymes, and compromised ATP generation (Shao, Chen et al. 2015).

It is important to emphasize that the selected metabolites or functional domains in our study were unaffected by MetS or BMI, as well as sex and age, and so represent a metabolomic profile indicative of fundamental pathophysiological processes in MDD rather than metabolic comorbidities. Even more, while no associations were detected among the metabolites and MetS, there were strong associations with MDD features. Consequently, the metabolic anomalies in MDD signify a non-MetS, immune–redox lipid-remodeling phenotype in MDD.

### Functional Metabotyping in MDD

The functional pattern of the metabolites determined in this study suggests a convergence of diaglycerol (DAG) - protein kinase C (PKC) lipotoxicity, membrane phospholipid remodeling, mitochondrial redox overload, ether-lipid depletion, and retinoid deficiency, all of which are integral to the NIMETOX model of depression (Maes, Almulla et al. 2025). Initially, elevations in 1-stearoyl-2-arachidonoylglycerol and myristic acid, alongside reductions in DG(18:1/18:2), characterize a transition toward pro-inflammatory DAG variants that stimulate PKC, hinder endothelial signaling, and enhance ROS generation (Samuel and Shulman 2012). A reduction in DG(18:1/18:2) signifies a depletion of a comparatively “neutral” unsaturated DAG pool, perhaps leading to an increase in the relative proportion of pro-inflammatory DAGs like 18:0/20:4. These AA/saturated-enriched DAGs are effective PKC activators, promoting NF-κB, cytokine synthesis, and lipotoxic signaling. The transition from neutral to bioactive diacylglycerol species is an acknowledged process in lipotoxic inflammation (Samuel and Shulman, 2012).

Secondly, our observations indicate that elevated methyl stearate, along with diminished arachidic acid (C20:0), implies a disruption in the typical processing of saturated fatty acids, resulting in an excess of stearate in fatty acid methyl esters. Elevated methyl stearate may indicate a detoxification or overflow mechanism that is activated when mitochondrial or peroxisomal β-oxidation are saturated or compromised (Houten and Wanders 2010, Rui 2014). A decline in arachidic acid signifies a reduction in relatively inert saturated storage pools, potentially shifting fatty acid metabolism toward more unsaturated, bioactive species that facilitate lipotoxic pathways (Tvrdik, Westerberg et al. 2000). Consequently, reduced levels of C20:0 indicate a shift from secure storage to active modification of fatty acids.

Third, a predominant cluster comprises enhanced phospholipids, including PI(22:4/16:0), PS(22:5/22:4), PS(22:5ω3/22:4), and PC(18:0/20:4). These molecules constitute the metabolic substrate for lipoxygenase activation, facilitating subsequent leukotriene and eoxin synthesis, as demonstrated by the elevation of 14,15-leukotriene C4 or eoxinC4, a powerful mediator of vascular inflammation (Feltenmark et al., 2008). These phospholipids undergo preferential oxidation, resulting in endothelial injury, phosphatidylserine externalization, microparticle release, and blood-brain barrier breakdown (Gianazza, Brioschi et al. 2021). The elevation observed in 1,2-dioleoyl-PI likely reflects increased turnover of the PI cycle, thereby promoting DAG generation and PKC activation (Berridge and Irvine 1989, Nishizuka 1992). Increased eoxin C4 production, membrane remodeling, and depletion of ether-lysophosphocholine may all point to an involvement of activated phospholipase A_2_ (PLA_2_) and/or increased immune-redox stress (Kudo and Murakami, 2002).

The mitochondrial–redox zone within our panel is distinguished by hydroxyphenyllactic acid, cysteinylhistidine, and inosinic acid. Hydroxyphenyllactic acid reflects the metabolism of aromatic amino acids (tyrosine) and intestinal microbiota during systemic stress and redox imbalances (Chernevskaya, Beloborodova et al. 2020). Cysteylhistidine formation may be the consequence of protein thiol oxidation and disulfide formation, corresponding with increased levels of ROS/RNS within the NIMETOX framework (Maes, Galecki et al. 2011). Inosinic acid (IMP), a key metabolite resulting from the degradation of ATP/AMP, is elevated following mitochondrial energy failure or inflammatory stress, reflecting disrupted purine metabolism (Harkness 1988, Houten and Wanders 2010). Together, these three metabolites define a unified axis of mitochondrial dysfunction, redox impairments, and gut-derived aromatic stress.

The decrease of retinyl-acetate glucuronide indicates aberrations in vitamin A metabolism or retinoid depletion resulting from extended oxidative detoxification (Penniston and Tanumihardjo 2005). This aligns with lipid peroxidation and diminished antioxidant capacity in MDD (Maes, Almulla et al. 2025). The reduction of 1-O-hexadecyl-lysoPC reflects a depletion of ether-lipids as a consequence of immune-redox stress (Stafforini 2009).

Simultaneous elevations of tetranor-12(S)-HETE, PI3P (1-phosphatidyl-1D-myo-inositol 3-phosphate, PI3P), and cysteylhistidine in patients with increasing ROI likely reflect a shared pathway of chronic membrane-redox stress and inflammatory lipid signaling. Tetranor-12(S)-HETE, the β-oxidation end product of lipoxygenase-12-derived 12-HETE, reflects extended turnover of arachidonic and adrenic eicosanoids and the generation of reactive oxygen species (Kühn and O’Donnell 2006). Elevated PI3P levels may indicate an accelerated phosphatidylinositol cycle and, consequently, enhanced endosomal and TLR signaling (Berridge and Irvine 1989). Both markers, in conjunction with elevated cysteinylhistidine, may form a self-sustaining lipid–redox–inflammatory cycle that perpetuates cellular stress, thereby increasing the susceptibility to increasing ROI and NIMETOX pathways. Previously, it was established that ROI is associated with many NIMETOX pathways including increased atherogenicity (Maes, Almulla et al. 2025).

### Metabolomics-Driven Diagnostic Precision

Through a meticulously regulated multistage machine-learning architecture, we found a 16-metabolite panel that effectively differentiated MDD from controls across Neural Networks, Linear Discriminant Analysis, and Support Vector Machines. The findings are bolstered by rigorous compliance with leakage-prevention protocols: all variable selection, preprocessing, model training, and tuning were restricted to the training sample, while performance assessment was conducted solely on testing or holdout samples, thereby ensuring unbiased estimates and preventing model overfitting.

Employing PLS-derived case-wise contribution scores, we demonstrated that each patient possesses a unique configuration of the 16 metabolites and six lipid–redox dimensions, despite comparable total depression ratings. This suggests that MDD is not a single metabolic or NIMETOX entity but rather a spectrum of partially overlapping metabotypes. Individualized profiles can guide mechanism-based treatment choices, favoring anti-inflammatory or lipoxygenase-12/15 targeted therapies in cases with high PL remodeling or lipotoxicity, and mitochondrial support in instances characterized by low ether-lipids and redox-driven pathology. This corresponds with the principle of nomothetic precision psychiatry wherein biological dimensions, rather than syndromic classifications, dictate treatment selection and sequencing (Maes, Moraes et al. 2023, Maes, Zhou et al. 2024),

Furthermore, the incorporation of metabolomic scores with clinical data and additional biomarkers (inflammation, HDL/ paraoxonase 1, PON1, genetics, microbiota) may facilitate multimodal categorization into physiologically coherent groupings, as opposed to employing trial-and-error medication. Future research may evaluate therapy algorithms that randomize individuals based not alone on illness, but also on metabolic phenotype (e.g., “lipotoxic–mitochondrial” versus “inflammatory–membrane remodeling”). Such designs would allow for the assessment of whether aligning medications with the predominant lipid–redox pathophysiology improves response rates and decreases non-response in comparison to standard care (Almulla and Maes 2025).

Although the results indicate a robust biological signal, such high precision scores must be regarded with caution and validated in larger and independent datasets (see below: limitations). Ultimately, translating these metabotypes into clinically applicable decision algorithms will be essential for progressing from descriptive metabolomics to true precision medicine in MDD.

### Upstream Pathways that may shape the MDD Metabotype

This section will address how the metabolites selected here may serve as downstream integrators of the upstream NIMETOX factors implicated in the pathophysiology of MDD, namely immune activation, glucocorticoid signals, oxidative/nitrosative stress, mitochondrial dysfunction, redox overload, endothelial activation, and compromised HDL/PON1-mediated detoxification (Maes, Almulla et al. 2025, Maes, Jirakran et al. 2025).

Hypercortisolemia and overactivity of the sympatho-adrenal system (SAS), both elevated in MDD (Maes, Meltzer et al. 1993, Carroll, Iranmanesh et al. 2012), serve as early catalysts. Catecholamines and cortisol augment whole-body lipolysis and fatty acid flow, which may increase 1-stearoyl-2-arachidonoylglycerol and myristic acid while diminishing benign oleic/linoleic diacylglycerol species such as DG(18:1/18:2) (Samuel and Shulman 2012). The outcome is a transition towards pro-inflammatory, arachidonate-rich DAGs, which activate PKC and produce mitochondrial ROS (see previous sections). Moreover, glucocorticoids, including cortisol, might compromise mitochondrial function and diminish the efficiency of oxidative phosphorylation (Du, Wang et al. 2009), potentially leading to ATP degradation and modified purine metabolism.

Elevations in ROS/RNS are a characteristic feature of the NIMETOX pathways in MDD (Maes, Galecki et al. 2011). Phospholipids with a high propensity for peroxidation, such as PI(22:4/16:0) and PS(22:5/22:4), are initial targets of ROS, leading to the exposure of PL oxidation, the production of microparticles, and increased susceptibility of the blood-brain barrier (Davies and Guo 2014). ROS could also interfere with retinoid turnover leading to diminished levels of retinyl-acetate glucuronide (Penniston and Tanumihardjo 2006). Impaired reverse cholesterol transport (RCT) and diminished antioxidant defenses, characterized by reduced HDL, decreased Apolipoprotein A1, and diminished PON1 activity, which are MDD biomarkers (Maes, Almulla et al. 2025, Maes, Jirakran et al. 2025), might compromise the clearance of oxidized phospholipids (Almulla, Thipakorn et al. 2023). Low levels of HDL and PON1 in MDD (Morris, Puri et al. 2021, Almulla, Thipakorn et al. 2023), may lead to the accumulation of oxidized arachidonic acid / adrenic phospholipids, hence elevating PS(22:5/22:4), PC(18:0/20:4), and PI(22:4/16:0). These data align with reduced levels of 1-O-hexadecyl-lysoPC, suggesting a reduction in endogenous ether-lipids as a consequence of oxidative stress and immune activation (Human Metabolome, 2025; HMDB, 2025).

The negative acute-phase inflammatory response, characterized by decreased albumin and transferrin, is another feature of MDD (Maes 1993). Reduced albumin levels diminish the buffering capacity of free fatty acids and oxidized phospholipids (van der Vusse 2009), likely promoting their integration into arachidonic acid-rich phosphatidylinositol / phosphatidylserine / phosphatidylcholine pools. Reduced transferrin levels elevate non-transferrin-bound iron, hence augmenting Fenton chemistry, and the peroxidation of long-chain polyunsaturated fatty acid phospholipids (Maes, Bosmans et al. 1997, Papanikolaou and Pantopoulos 2005). Ultimately, leaky gut and bacterial translocation (Maes, Kubera et al. 2013) may exacerbate lipid remodeling. LPS and bacterial translocation stimulate Toll-like receptor 4 – NF-κB and lipoxygenase-12/15 signaling pathways, resulting in elevated cytokine levels and ROS that may exacerbate PL remodeling (Balestrieri, Di Costanzo et al. 2021). In this respect, increased eoxin C4 production via 15-lipoxygenase is associated with endothelial inflammation and vascular permeability (Feltenmark et al., 2008).

Therefore, the metabolite signature identified herein embodies a cohesive metabolic network that connects the established pathophysiology of MDD to PL remodeling, lipotoxicity, eoxin C4 surplus, and ether-lipid depletion. These alterations may inform the brain and impact limbic-prefrontal circuits, resulting in affective symptoms (Maes, Almulla et al., 2025). Peripheral NIMETOX modules interact with brain functions through BBB disruption (initiated by eicosanoids, ROS, cytokines, etc.), endothelial and perivascular signaling (mediated by LOX and TLRs), ROS and cytokine production (triggered by LOX, DAG-PKC, PLA2), the vagal pathways (triggered by eicosanoids and cytokines), and shifts in energy metabolism (Maes, Almulla et al., 2025; Maes, Almulla et al., 2025).

Limitations

Some methodological constraints must be recognized. Although the sample of 125 MDD patients and 40 controls is larger than many prior metabolomic research, it is still modest for high-dimensional omics investigations, potentially restricting the generalizability of the detected metabolic biosignature. Second, despite meticulous data curation and the elimination of non-endogenous metabolites, untargeted metabolomics continues to be susceptible to batch effects, ion suppression, and platform-specific detection biases, potentially affecting quantitative precision. Third, the design is cross-sectional, which precludes causal inference. Fourth, despite the statistical correction of possible confounders (age, sex, BMI, metabolic syndrome, drug state of the patients), unmeasured variables such as food, circadian rhythms, physical activity, and gut microbiota composition may have influenced the metabolic variance. Fifth, the work concentrates solely on serum metabolomics, while the incorporation of concurrent transcriptome, proteomic, lipidomic, and microbiome data would yield a more comprehensive systems-biological perspective of NIMETOX processes in MDD. Sixth, although the present use of psychotropic medications did not influence the outcomes of this study, subsequent research should investigate the effects of psychotropics on the metabolites analyzed in this study. Despite the validation of various machine-learning methodologies (PLS-DA, LDA, NN, SVM) in testing and holdout samples with safeguards against data leakage, external replication in independent cohorts—encompassing diverse ethnicities, clinical environments, or medication statuses—is necessary to confirm genuine reproducibility.

## Conclusions

This study illustrates that serum metabolomics may detect a distinctive metabolite lipid–redox biosignature that accurately defines MDD and the severity of its phenome, namely OSOD, physiosomatic symptoms, suicidal ideation, and ROI. The pathway signature includes PL modifications, DAG–PKC lipotoxicity, ether-lipid depletion, mitochondrial redox stress, eoxin formation, and impaired retinoid detoxification, forming a cohesive metabolic framework consistent with the NIMETOX model of MDD. These findings highlight metabolomics as a comprehensive framework for biological subtyping and precision psychiatry, progressing from symptom-based diagnosis to mechanistically informed classification. We now perform external validation in different subtypes of MDD, namely first episode mild MDD in adults and adolescents, and an independent cohort from a different clinical site in another province in China to enhance generalizability and translational usefulness.

The uncovered pathways emphasize novel therapeutic targets, including lipoxygenase-15 signaling inhibitors, PLA_2_ inhibitors, agents that restore PL remodeling, enhancers of RCT, mitochondrial protectants such as CoQ10 and N-Acetyl-cysteine, and anti-inflammatory, pro-resolving lipid mediators (e.g. resolvins). Future investigations should assess whether modifications of these pathways yield clinical benefits. External validation and longitudinal studies are essential for evaluating whether the constructed metabolite panel and pathway signatures can accurately predict therapeutic response.

## Supporting information

Electronic Supplementary File

## Ethics approval

This study received approval from the ethics committee of Sichuan Provincial People’s Hospital [Ethics (Research) 2024-203] and was conducted in precise accordance with ethical standards and privacy regulations.

## Consent to participate

Prior to participating in this investigation, each participant provided written informed consent.

## Consent for publication

All authors have provided their consent for the publication of this paper.

## Author’s contributions

Michael Maes: supervision, conceptualization, formal analysis, writing - review and editing. Mengqi Niu: visualization, writing recruiting participants. Chenkai Yangyang and Jing Li: recruiting participants. Abbas F Almulla: editing, visualization; Yingqian Zhang: conceptualization, visualization, writing, review, editing.

## Data access statement

The database compiled during this study will be made available by the corresponding author (MM) upon a reasonable request, following the authors’ comprehensive utilization of the data set.

## References

Alberti, K. G., R. H. Eckel, S. M. Grundy, P. Z. Zimmet, J. I. Cleeman, K. A. Donato, J. C. Fruchart, W. P. James, C. M. Loria and S. C. Smith, Jr. (2009). “Harmonizing the metabolic syndrome: a joint interim statement of the International Diabetes Federation Task Force on Epidemiology and Prevention; National Heart, Lung, and Blood Institute; American Heart Association; World Heart Federation; International Atherosclerosis Society; and International Association for the Study of Obesity.” Circulation 120(16): 1640–1645.

Almulla, A. F. and M. Maes (2025). “Peripheral Immune-Inflammatory Pathways in Major Depressive Disorder, Bipolar Disorder, and Schizophrenia: Exploring Their Potential as Treatment Targets.” CNS Drugs 39(8): 739–762.

Almulla, A. F., Y. Thipakorn, A. A. A. Algon, C. Tunvirachaisakul, H. K. Al-Hakeim and M. Maes (2023). “Reverse cholesterol transport and lipid peroxidation biomarkers in major depression and bipolar disorder: A systematic review and meta-analysis.” Brain Behav Immun 113: 374–388.

Balestrieri, B., D. Di Costanzo and D. F. Dwyer (2021). “Macrophage-Mediated Immune Responses: From Fatty Acids to Oxylipins.” Molecules 27(1).

Beck, A. T., R. A. Steer and G. K. Brown (1996). BDI-II, Beck depression inventory: manual. San Antonio, Tex., Boston, Psychological Corp.; Harcourt Brace San Antonio, Tex., Boston.

Berridge, M. J. and R. F. Irvine (1989). “Inositol phosphates and cell signalling.” Nature 341(6239): 197–205.

Cai, H., T. Cao, N. Li, P. Fang, P. Xu, X. Wu, B. Zhang and D. Xiang (2019). “Quantitative monitoring of a panel of stress-induced biomarkers in human plasma by liquid chromatography-tandem mass spectrometry: an application in a comparative study between depressive patients and healthy subjects.” Anal Bioanal Chem 411(22): 5765–5777.

Carroll, B. J., A. Iranmanesh, D. M. Keenan, F. Cassidy, W. H. Wilson and J. D. Veldhuis (2012). “Pathophysiology of hypercortisolism in depression: pituitary and adrenal responses to low glucocorticoid feedback.” Acta Psychiatr Scand 125(6): 478–491.

Chan, P., I. Suridjan, D. Mohammad, N. Herrmann, G. Mazereeuw, L. M. Hillyer, D. W. L. Ma, P. I. Oh and K. L. Lanctôt (2018). “Novel Phospholipid Signature of Depressive Symptoms in Patients With Coronary Artery Disease.” J Am Heart Assoc 7(10).

Chernevskaya, E., N. Beloborodova, N. Klimenko, A. Pautova, D. Shilkin, V. Gusarov and A. Tyakht (2020). “Serum and fecal profiles of aromatic microbial metabolites reflect gut microbiota disruption in critically ill patients: a prospective observational pilot study.” Crit Care 24(1): 312.

Davies, S. S. and L. Guo (2014). “Lipid peroxidation generates biologically active phospholipids including oxidatively N-modified phospholipids.” Chem Phys Lipids 181: 1–33.

Du, J., Y. Wang, R. Hunter, Y. Wei, R. Blumenthal, C. Falke, R. Khairova, R. Zhou, P. Yuan, R. Machado-Vieira, B. S. McEwen and H. K. Manji (2009). “Dynamic regulation of mitochondrial function by glucocorticoids.” Proc Natl Acad Sci U S A 106(9): 3543–3548.

Feltenmark, S., Gautam, N., Brunnström, A., Griffiths, W., Backman, L., Edenius, C., Lindbom, L., Björkholm, M., Claesson, H.E. (2008) Eoxins are proinflammatory arachidonic acid metabolites produced via the 15-lipoxygenase-1 pathway in human eosinophils and mast cells. Proc Natl Acad Sci U S A. 105(2):680–685.

Gianazza, E., M. Brioschi, A. Martinez Fernandez, F. Casalnuovo, A. Altomare, G. Aldini and C. Banfi (2021). “Lipid Peroxidation in Atherosclerotic Cardiovascular Diseases.” Antioxid Redox Signal 34(1): 49–98.

Gierk, B., S. Kohlmann, K. Kroenke, L. Spangenberg, M. Zenger, E. Brähler and B. Löwe (2014). “The somatic symptom scale-8 (SSS-8): a brief measure of somatic symptom burden.” JAMA Intern Med 174(3): 399–407.

Hamilton, M. (1959). “The assessment of anxiety states by rating.” British journal of medical psychology.

Hamilton, M. (1960). “A rating scale for depression.” Journal of neurology, neurosurgery, and psychiatry 23(1): 56.

Harkness, R. A. (1988). “Hypoxanthine, xanthine and uridine in body fluids, indicators of ATP depletion.” J Chromatogr 429: 255–278.

Houten, S. M. and R. J. Wanders (2010). “A general introduction to the biochemistry of mitochondrial fatty acid β-oxidation.” J Inherit Metab Dis 33(5): 469–477.

Kaddurah-Daouk, R., S. H. Boyle, W. Matson, S. Sharma, S. Matson, H. Zhu, M. B. Bogdanov, E. Churchill, R. R. Krishnan, A. J. Rush, E. Pickering and M. Delnomdedieu (2011). “Pretreatment metabotype as a predictor of response to sertraline or placebo in depressed outpatients: a proof of concept.” Transl Psychiatry 1(7): e26-.

Kim, E. Y., J. W. Lee, M. Y. Lee, S. H. Kim, H. J. Mok, K. Ha, Y. M. Ahn and K. P. Kim (2018). “Serum lipidomic analysis for the discovery of biomarkers for major depressive disorder in drug-free patients.” Psychiatry Res 265: 174–182.

Kudo, I., Murakami, M. (2002) Phospholipase A2 enzymes. Prostaglandins Other Lipid Mediat. 68–69:3-58.

Kühn, H. and V. B. O’Donnell (2006). “Inflammation and immune regulation by 12/15-lipoxygenases.” Prog Lipid Res 45(4): 334–356.

Kuhn, M. and K. Johnson (2013). Applied predictive modeling, Springer.

Leonard, B. and M. Maes (2012). “Mechanistic explanations how cell-mediated immune activation, inflammation and oxidative and nitrosative stress pathways and their sequels and concomitants play a role in the pathophysiology of unipolar depression.” Neurosci Biobehav Rev 36(2): 764–785.

Leroy, L. A., A. Mac Donald, A. Kandlur, D. Bose, P. Xiao, J. Gagnon, F. Villinger and Y. Chebloune (2022). “Cytokine Adjuvants IL-7 and IL-15 Improve Humoral Responses of a SHIV LentiDNA Vaccine in Animal Models.” Vaccines (Basel) 10(3).

Liu, Y., L. Yieh, T. Yang, W. Drinkenburg, P. Peeters, T. Steckler, V. A. Narayan, G. Wittenberg and J. Ye (2016). “Metabolomic biosignature differentiates melancholic depressive patients from healthy controls.” BMC Genomics 17(1): 669.

Maes, M. (1993). “A review on the acute phase response in major depression.” Rev Neurosci 4(4): 407–416.

Maes, M. (2022). “Precision Nomothetic Medicine in Depression Research: A New Depression Model, and New Endophenotype Classes and Pathway Phenotypes, and A Digital Self.” J Pers Med 12(3).

Maes, M., A. F. Almulla, S. Drozdstoj and Y. Zhang (2025). “Hallmarks of major depression: neuroimmune-metabolic-oxidative (NIMETOX) pathways.” ResearchGate.

Maes, M., A. F. Almulla, Z. You and Y. Zhang (2025). “Neuroimmune, metabolic and oxidative stress pathways in major depressive disorder.” Nature Reviews Neurology 21(9): 473–489.

Maes, M., E. Bosmans, S. Scharpé, D. Hendriks, W. Cooremans, H. Neels, F. De Meyer, P. D’Hondt and D. Peeters (1997). “Components of biological variation in serum soluble transferrin receptor: relationships to serum iron, transferrin and ferritin concentrations, and immune and haematological variables.” Scand J Clin Lab Invest 57(1): 31–41.

Maes, M., P. Galecki, Y. S. Chang and M. Berk (2011). “A review on the oxidative and nitrosative stress (O&NS) pathways in major depression and their possible contribution to the (neuro)degenerative processes in that illness.” Prog Neuropsychopharmacol Biol Psychiatry 35(3): 676–692.

Maes, M., K. Jirakran, L. O. Semeão, A. P. Michelin, A. K. Matsumoto, F. F. Brinholi, D. S. Barbosa, C. Tivirachaisakul, A. F. Almulla, D. Stoyanov and Y. Zhang (2025). “Key factors underpinning neuroimmune-metabolic-oxidative (NIMETOX) major depression in outpatients: paraoxonase 1 activity, reverse cholesterol transport, increased atherogenicity, protein oxidation, and differently expressed cytokine networks.” Neuro Endocrinol Lett 46(2): 115–125.

Maes, M., K. Jirakran, A. Vasupanrajit, M. Niu, B. Zhou, D. S. Stoyanov and C. Tunvirachaisakul (2024). “The recurrence of illness (ROI) index is a key factor in major depression that indicates increasing immune-linked neurotoxicity and vulnerability to suicidal behaviors.” Psychiatry Res 339: 116085.

Maes, M., M. Kubera, J. C. Leunis, M. Berk, M. Geffard and E. Bosmans (2013). “In depression, bacterial translocation may drive inflammatory responses, oxidative and nitrosative stress (O&NS), and autoimmune responses directed against O&NS-damaged neoepitopes.” Acta Psychiatrica Scandinavica 127(5): 344–354.

Maes, M., H. Y. Meltzer, E. Suy, B. Minner, J. Calabrese and P. Cosyns (1993). “Sleep disorders and anxiety as symptom profiles of sympathoadrenal system hyperactivity in major depression.” J Affect Disord 27(3): 197–207.

Maes, M., J. B. Moraes, A. Congio, H. Vargas and S. Nunes (2023). “Research and Diagnostic Algorithmic Rules (RADAR) for mood disorders, recurrence of illness, suicidal behaviours, and the patient’s lifetime trajectory.” Acta Neuropsychiatr 35(2): 104–117.

Maes, M., B. Zhou, K. Jirakran, A. Vasupanrajit, P. Boonchaya-Anant, C. Tunvirachaisakul, X. Tang, J. Li and A. F. Almulla (2024). “Towards a major methodological shift in depression research by assessing continuous scores of recurrence of illness, lifetime and current suicidal behaviors and phenome features.” J Affect Disord 350: 728–740.

Maes, M., A. F. Almulla, S. Drozdstoj and Y. Zhan (2025). “Advancements in the molecular understanding of major depressive disorder uncovering novel targets with therapeutic promise: focus on recurrence of illness.” ResearchGate. DOI: 10.13140/RG.2.2.23656.12803

Maes, M., A. F. Almulla, S. Drozdstoj, and Y. Zhan (2025). Hallmarks of major depression: neuroimmune-metabolic-oxidative (NIMETOX) pathways. ReserachGate, DOI: 10.13140/RG.2.2.22494.50244

Morris, G. and M. Maes (2014). “Oxidative and Nitrosative Stress and Immune-Inflammatory Pathways in Patients with Myalgic Encephalomyelitis (ME)/Chronic Fatigue Syndrome (CFS).” Curr Neuropharmacol 12(2): 168–185.

Morris, G., B. K. Puri, C. C. Bortolasci, A. Carvalho, M. Berk, K. Walder, E. G. Moreira and M. Maes (2021). “The role of high-density lipoprotein cholesterol, apolipoprotein A and paraoxonase-1 in the pathophysiology of neuroprogressive disorders.” Neurosci Biobehav Rev 125: 244–263.

Nishizuka, Y. (1992). “Intracellular signaling by hydrolysis of phospholipids and activation of protein kinase C.” Science 258(5082): 607–614.

Niu, M., Y. Zhang, X. Zhang, C. Yangyang, X. Zhuang, J. Li and M. Maes (2025). “Dissection of the clinical phenome of major depressive disorder into subdomains and subgroups in relation to activated immune-inflammatory profiles.” medRxiv: 2025.2010.2021.25338439.

Papanikolaou, G. and K. Pantopoulos (2005). “Iron metabolism and toxicity.” Toxicol Appl Pharmacol 202(2): 199–211.

Penniston, K. L. and S. A. Tanumihardjo (2005). “Elevated serum concentrations of beta-glucuronide metabolites and 4-oxoretinol in lactating sows after treatment with vitamin A: a model for evaluating supplementation in lactating women.” Am J Clin Nutr 81(4): 851–858.

Penniston, K. L. and S. A. Tanumihardjo (2006). “Vitamin A intake of captive rhesus monkeys exceeds national research council recommendations.” Am J Primatol 68(11): 1114–1119.

Posner, K., G. K. Brown, B. Stanley, D. A. Brent, K. V. Yershova, M. A. Oquendo, G. W. Currier, G. A. Melvin, L. Greenhill, S. Shen and J. J. Mann (2011). “The Columbia-Suicide Severity Rating Scale: initial validity and internal consistency findings from three multisite studies with adolescents and adults.” Am J Psychiatry 168(12): 1266–1277.

Rui, L. (2014). “Energy metabolism in the liver.” Compr Physiol 4(1): 177–197.

Samuel, V. T. and G. I. Shulman (2012). “Mechanisms for insulin resistance: common threads and missing links.” Cell 148(5): 852–871.

Shao, W. H., J. J. Chen, S. H. Fan, Y. Lei, H. B. Xu, J. Zhou, P. F. Cheng, Y. T. Yang, C. L. Rao, B. Wu, H. P. Liu and P. Xie (2015). “Combined Metabolomics and Proteomics Analysis of Major Depression in an Animal Model: Perturbed Energy Metabolism in the Chronic Mild Stressed Rat Cerebellum.” Omics 19(7): 383–392.

Somani, A., A. K. Singh, B. Gupta, S. Nagarkoti, P. K. Dalal and M. Dikshit (2022). “Oxidative and Nitrosative Stress in Major Depressive Disorder: A Case Control Study.” Brain Sci 12(2).

Spielberger, C., R. Gorsuch, R. Lushene, P. Vagg and G. Jacobs (1983). “Manual for the State-Trait Anxiety Inventory; Palo Alto, CA, Ed.” Palo Alto: Spielberger.

Stafforini, D. M. (2009). “Biology of platelet-activating factor acetylhydrolase (PAF-AH, lipoprotein associated phospholipase A2).” Cardiovasc Drugs Ther 23(1): 73–83.

Tvrdik, P., R. Westerberg, S. Silve, A. Asadi, A. Jakobsson, B. Cannon, G. Loison and A. Jacobsson (2000). “Role of a new mammalian gene family in the biosynthesis of very long chain fatty acids and sphingolipids.” J Cell Biol 149(3): 707–718.

van der Vusse, G. J. (2009). “Albumin as fatty acid transporter.” Drug Metab Pharmacokinet 24(4): 300–307.

Vaváková, M., Z. Ďuračková and J. Trebatická (2015). “Markers of Oxidative Stress and Neuroprogression in Depression Disorder.” Oxid Med Cell Longev 2015: 898393.

Zachrisson, O., B. Regland, M. Jahreskog, M. Kron and C. G. Gottfries (2002). “A rating scale for fibromyalgia and chronic fatigue syndrome (the FibroFatigue scale).” J Psychosom Res 52(6): 501–509.

Zheng, P., J. J. Chen, T. Huang, M. J. Wang, Y. Wang, M. X. Dong, Y. J. Huang, L. K. Zhou and P. Xie (2013). “A novel urinary metabolite signature for diagnosing major depressive disorder.” J Proteome Res 12(12): 5904–5911.

