## Supplementary material for "Peripheral Metabolic-Redox Signaling as a Core Mechanism of Major Depressive Disorder: Evidence From Deep Metabolomic Phenotyping": Electronic Supplementary File

### Slide 1
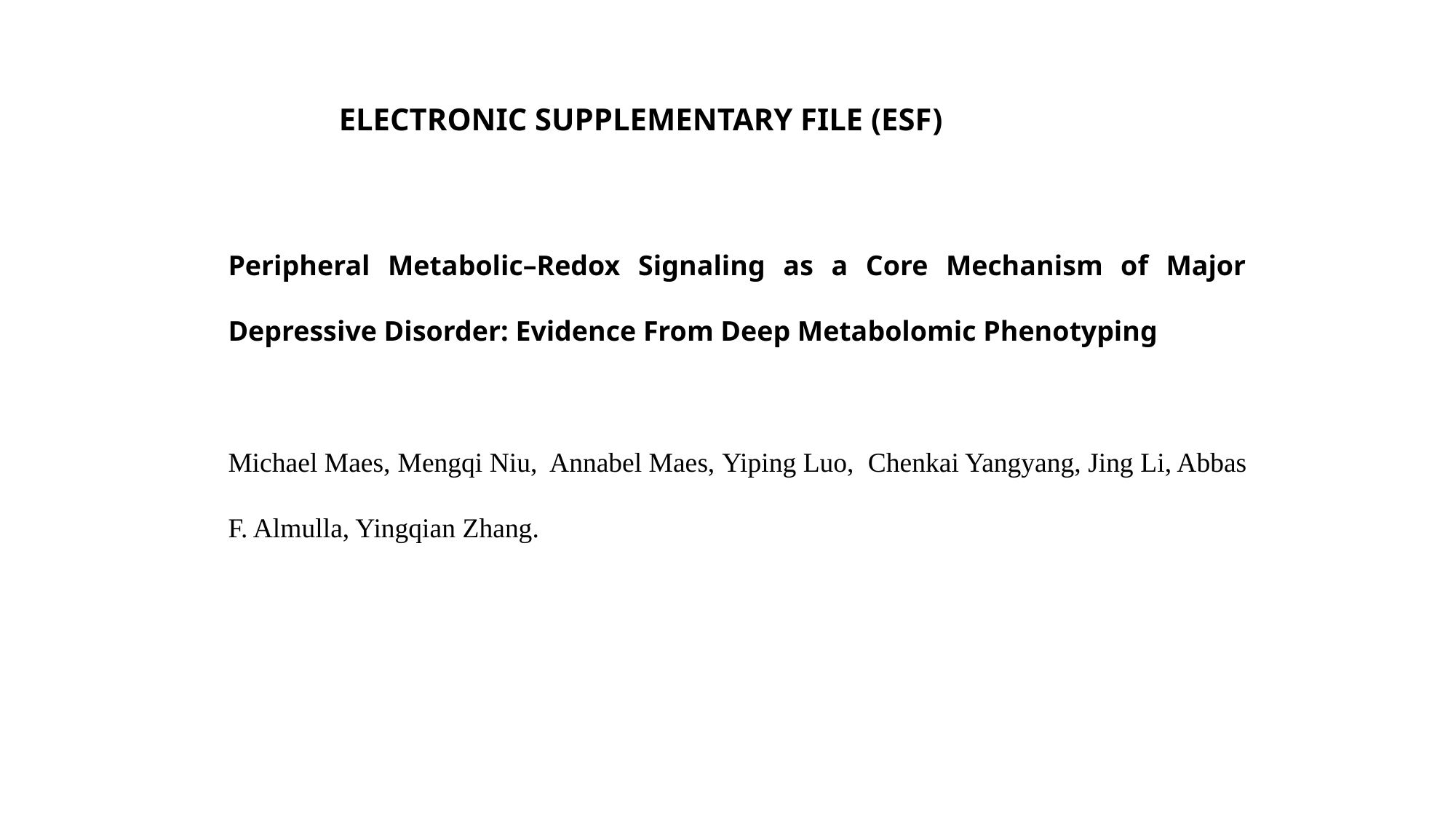

ELECTRONIC SUPPLEMENTARY FILE (ESF)
Peripheral Metabolic–Redox Signaling as a Core Mechanism of Major Depressive Disorder: Evidence From Deep Metabolomic Phenotyping
Michael Maes, Mengqi Niu, Annabel Maes, Yiping Luo, Chenkai Yangyang, Jing Li, Abbas F. Almulla, Yingqian Zhang.

### Slide 2
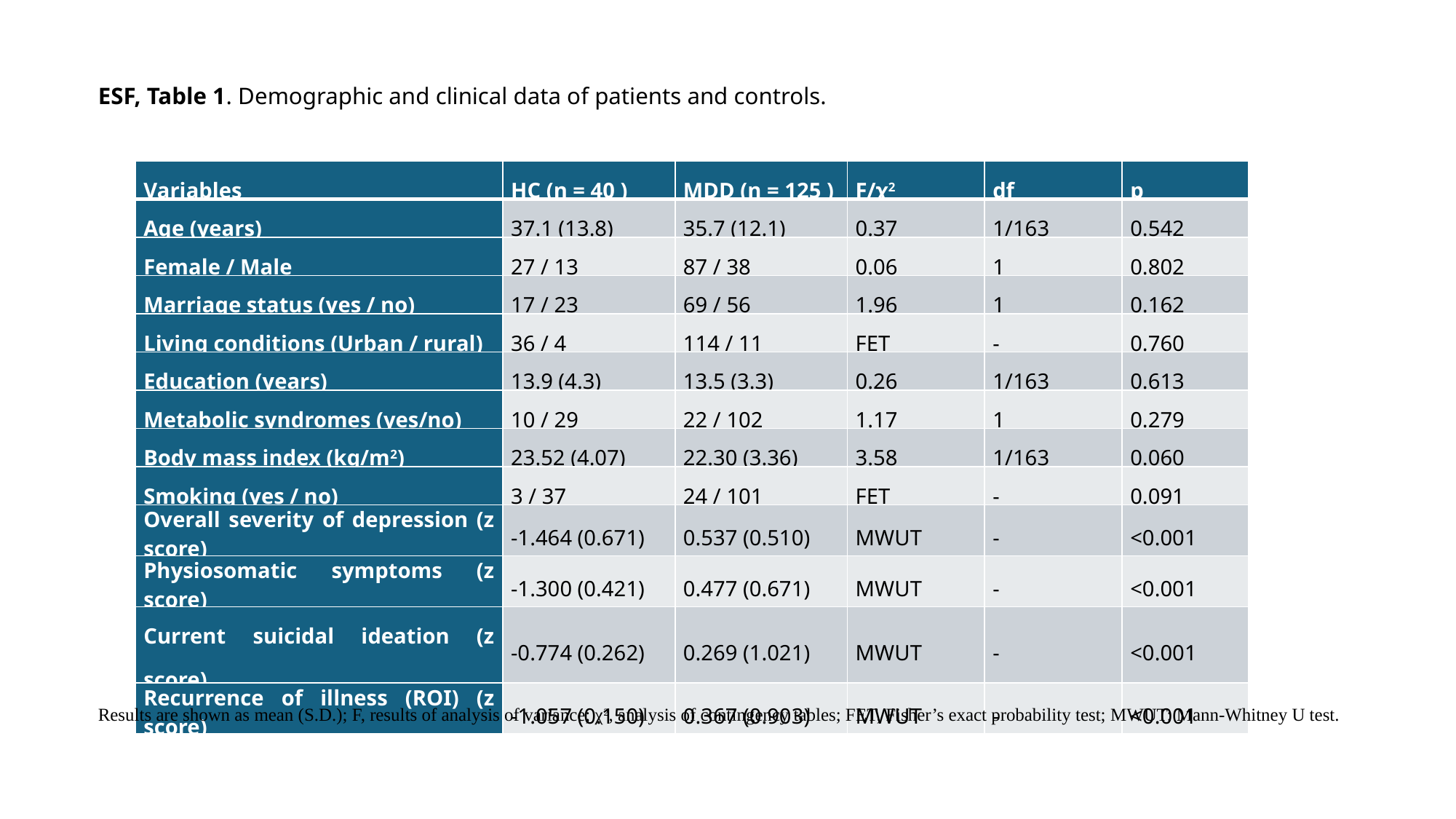

ESF, Table 1. Demographic and clinical data of patients and controls.
| Variables | HC (n = 40 ) | MDD (n = 125 ) | F/χ^2^ | df | p |
| --- | --- | --- | --- | --- | --- |
| Age (years) | 37.1 (13.8) | 35.7 (12.1) | 0.37 | 1/163 | 0.542 |
| Female / Male | 27 / 13 | 87 / 38 | 0.06 | 1 | 0.802 |
| Marriage status (yes / no) | 17 / 23 | 69 / 56 | 1.96 | 1 | 0.162 |
| Living conditions (Urban / rural) | 36 / 4 | 114 / 11 | FET | - | 0.760 |
| Education (years) | 13.9 (4.3) | 13.5 (3.3) | 0.26 | 1/163 | 0.613 |
| Metabolic syndromes (yes/no) | 10 / 29 | 22 / 102 | 1.17 | 1 | 0.279 |
| Body mass index (kg/m^2^) | 23.52 (4.07) | 22.30 (3.36) | 3.58 | 1/163 | 0.060 |
| Smoking (yes / no) | 3 / 37 | 24 / 101 | FET | - | 0.091 |
| Overall severity of depression (z score) | -1.464 (0.671) | 0.537 (0.510) | MWUT | - | <0.001 |
| Physiosomatic symptoms (z score) | -1.300 (0.421) | 0.477 (0.671) | MWUT | - | <0.001 |
| Current suicidal ideation (z score) | -0.774 (0.262) | 0.269 (1.021) | MWUT | - | <0.001 |
| Recurrence of illness (ROI) (z score) | -1.057 (0.150) | 0.367 (0.903) | MWUT | - | <0.001 |
Results are shown as mean (S.D.); F, results of analysis of variance; χ^2^, analysis of contingency tables; FET, Fisher’s exact probability test; MWUT: Mann-Whitney U test.

### Slide 3
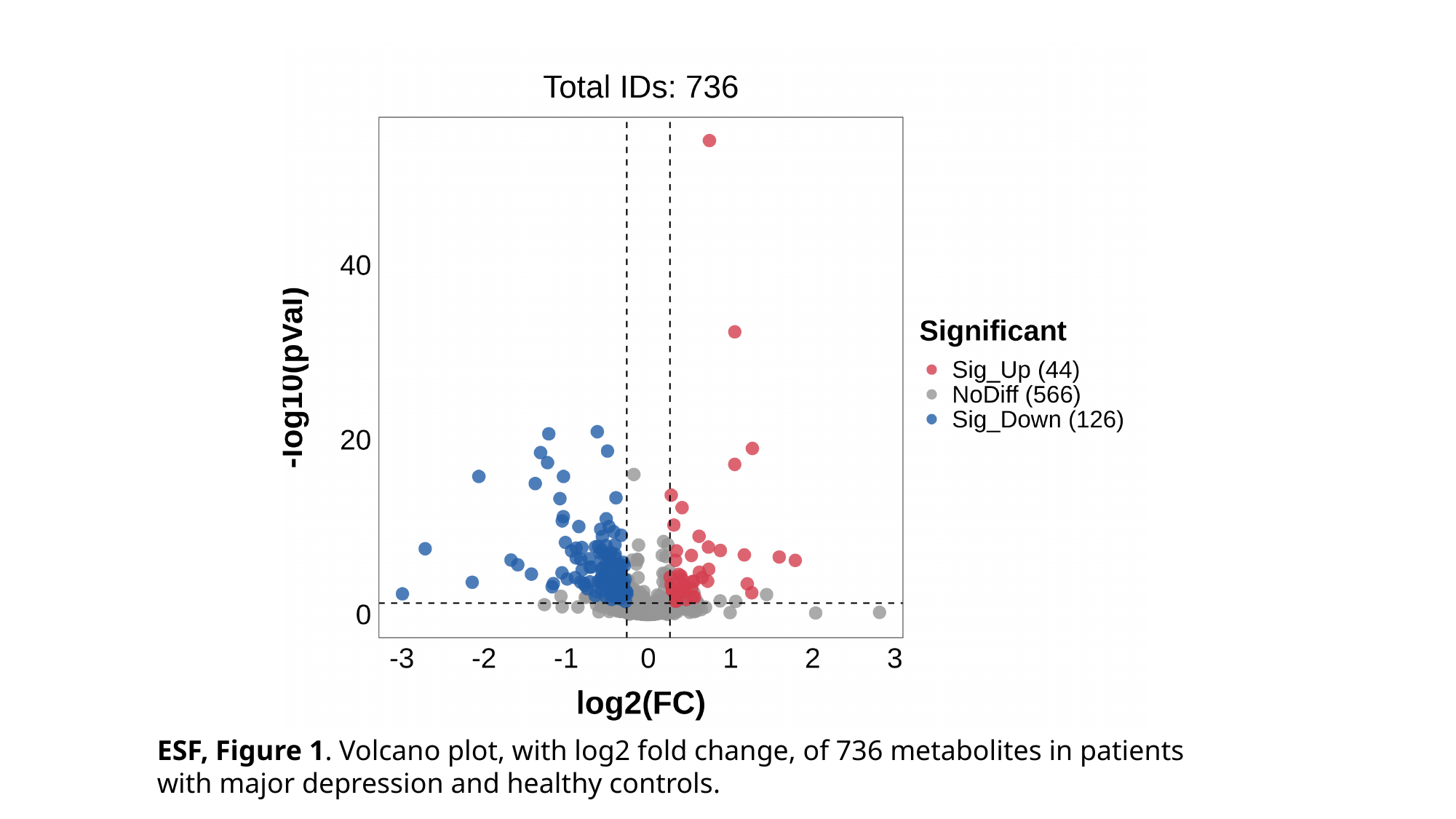

ESF, Figure 1. Volcano plot, with log2 fold change, of 736 metabolites in patients with major depression and healthy controls.

### Slide 4
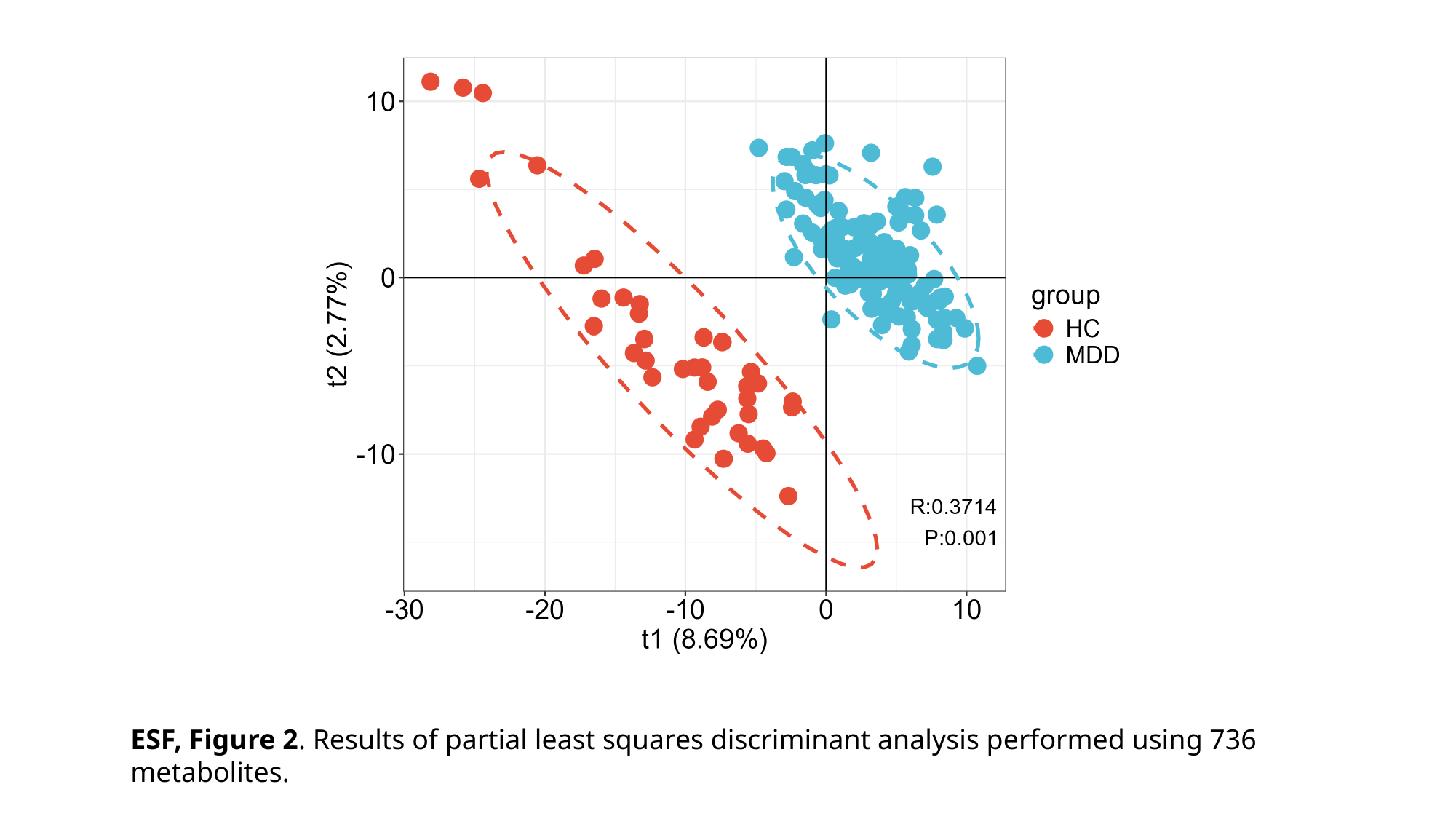

ESF, Figure 2. Results of partial least squares discriminant analysis performed using 736 metabolites.

### Slide 5
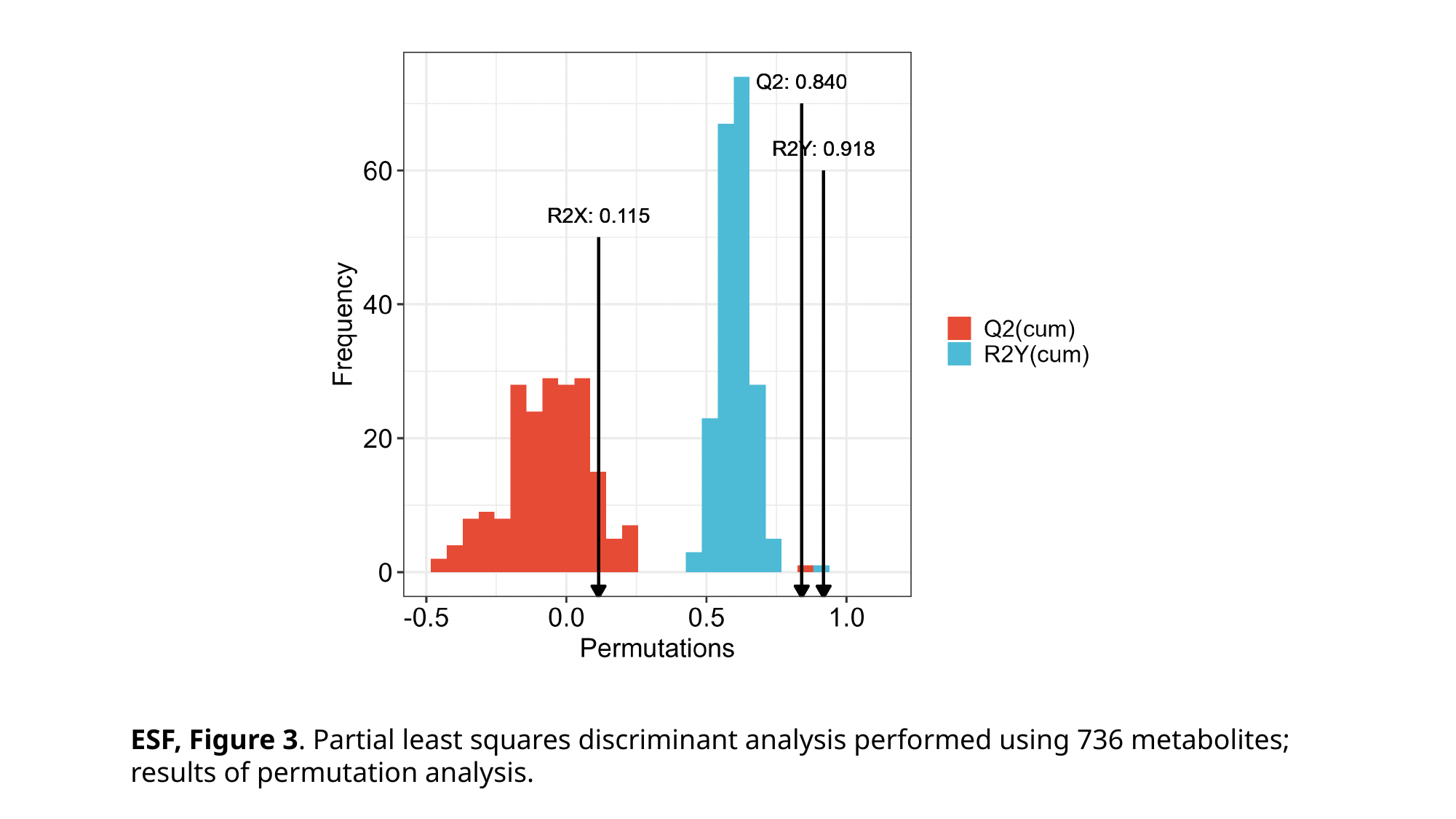

ESF, Figure 3. Partial least squares discriminant analysis performed using 736 metabolites; results of permutation analysis.

### Slide 6
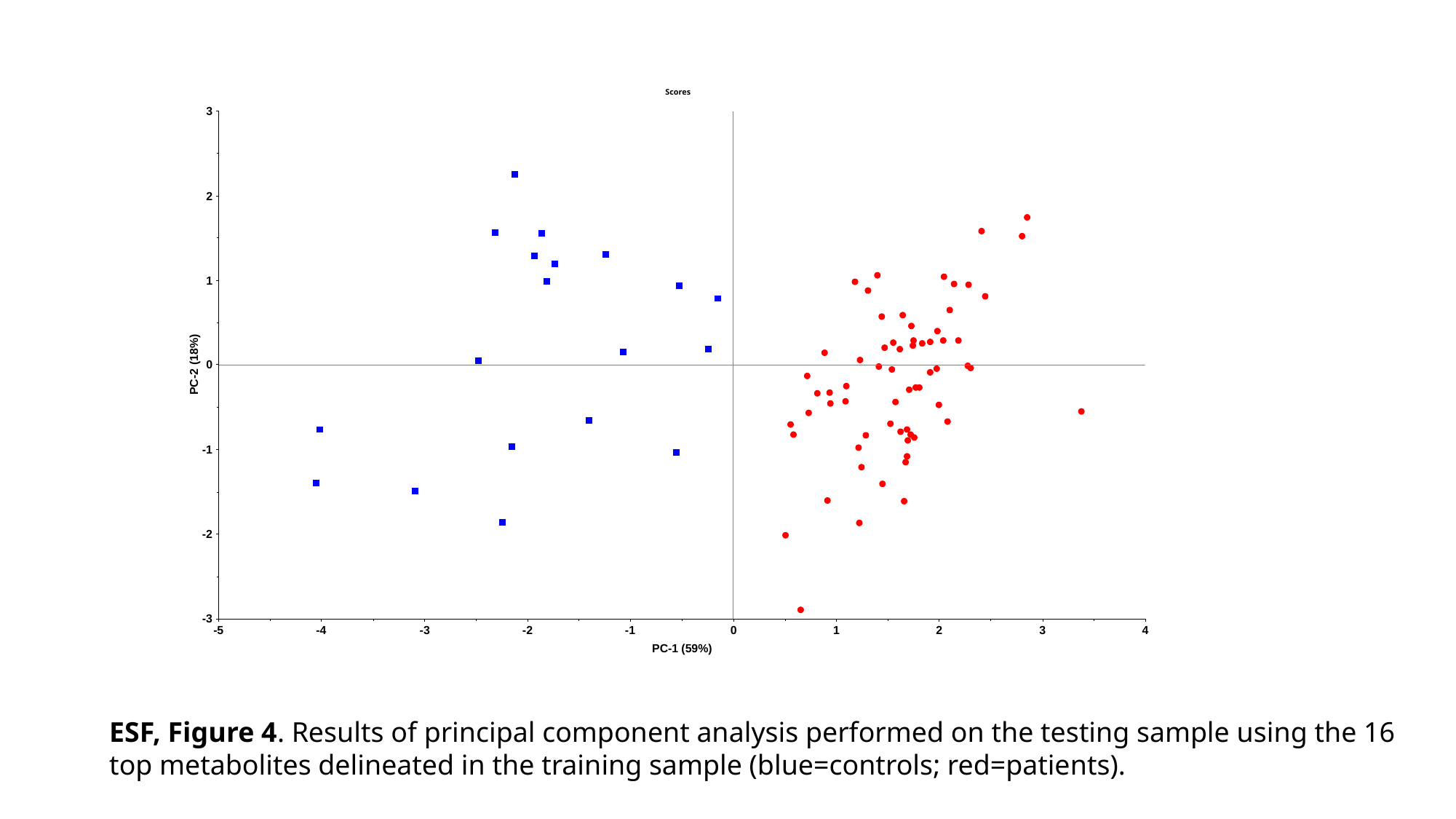

ESF, Figure 4. Results of principal component analysis performed on the testing sample using the 16 top metabolites delineated in the training sample (blue=controls; red=patients).
